# *TECTB* Variants Reveal Tectorial Membrane Vulnerability in Dominant Non-Syndromic Hearing Loss

**DOI:** 10.1101/2025.08.13.25333146

**Authors:** Evan B. Hale, Barbara Vona, Richard J. Goodyear, Richard T. Osgood, Jun Shen, Sami S. Amr, Karen Mojica, Michaela A. H. Hofrichter, Jörg Schröder, Sophie Flandin, Paul Gratias, Ricardo Vera-Monroy, Katherine Callahan, Kerry L. Gudlewski, Rolen Quadros, Masato Ohtsuka, JoAnn McGee, Edward J. Walsh, Cynthia C. Morton, Thomas Haaf, Channabasavaiah Gurumurthy, James E. Saunders, Guy P. Richardson, Artur A. Indzhykulian

## Abstract

Identifying new genes responsible for non-syndromic hearing loss remains a critical goal as many patients still lack a molecular diagnosis despite comprehensive genetic testing. The tectorial membrane (TM) is a specialized acellular matrix of the inner ear, essential for stimulating mechanosensitive hair cell stereocilia bundles and maintaining frequency tuning and auditory sensitivity. Although mutations in genes encoding several non-collagenous proteins found in the TM (TECTA, CEACAM16, OTOG, OTOGL) have been identified as deafness genes, definitive evidence implicating β-tectorin (TECTB) has been lacking.

Here, we present multiple lines of genetic and experimental evidence linking heterozygous missense variants in *TECTB* (NM_058222.3:c.674G>A, p.Cys225Tyr and NM_058222.3:c.853C>T, p.Arg285Cys), with hearing loss. Each variant affects highly conserved residues within or directly flanking the zona pellucida domain. Using a *Tectb-C225Y* knock-in mouse model, we show that homozygous animals exhibit severe hearing loss and profound disruption of TM morphology, while heterozygous animals display decreased staining density within the TM and increased susceptibility to noise-induced hearing loss, despite normal auditory thresholds.

These findings identify *TECTB* as a novel human deafness gene, further elucidate its contribution to maintaining TM integrity and resilience against environmentally-related auditory decline.

## Introduction

Hearing loss is the most prevalent human sensory deficit (Armstrong et al., 2022) and can result from disruptions at any point along the auditory pathway. Amplification and transduction of sound stimuli are mediated by hair cells, the mechanosensory cells of the inner ear (Corey & Hudspeth, 1979; Robles & Ruggero, 2001). The hair cells are arrayed along the tonotopic axis of the cochlea and specific regions are tuned to respond best to different sound frequencies.

Housed between the basilar and tectorial membranes (TM) and located within the organ of Corti, the hair cells respond locally to frequency-specific mechanical stimuli. The outer hair cells facilitate amplification of sound-induced basilar membrane motion through electromotility – rapid cycles of contraction and elongation – while inner hair cells (IHCs) convert mechanical stimuli into neural signals that are transmitted via the auditory nerve to the brain (Dallos, 2008). The defining feature of hair cells is a bundle of stereocilia, a set of actin-rich hair-like projections on the apical surface of each cell. Bundle deflection generates tension in links interconnecting the stereocilia, thereby opening mechanosensitive ion channels at the tips of all but the tallest stereocilia (Corey & Hudspeth, 1979; Fettiplace & Kim, 2014; Robles & Ruggero, 2001). The TM shears relative to the apical surface of the organ of Corti in response to sound waves, applying force to the hair bundles and facilitating mechanoelectrical transduction (Hakizimana & Fridberger, 2021; Verpy et al., 2011). The structural integrity of the TM is therefore critical for cochlear sensitivity, frequency discrimination – the sharpness of tuning along the tonotopic axis – and gain (Legan et al., 2000).

The TM is a specialized structure essential for auditory transduction that is composed, in mammals, of collagen fibers that are organized by and embedded in a matrix formed by a number of non-collagenous glycoproteins including CEACAM16, α-tectorin (TECTA) and ꞵ-tectorin (TECTB) (Goodyear & Richardson, 2018). These proteins interact to form the striated-sheet matrix (SSM), an acellular structure critical for effectively coupling mechanical sound stimulation to hair cell stereocilia deflection (Goodyear & Richardson, 2002). Among them, TECTA is the largest and most structurally complex, with a predicted molecular mass of ∼239 kDa and 33 potential sites for N-glycosylation, while TECTB is smaller (∼36 kDa) with just 4 potential N-glycosylation sites. Despite these differences, TECTA and TECTB both contain a zona pellucida (ZP) domain (Jovine et al., 2005), a domain primarily defined by eight highly conserved cysteine residues that form intrachain disulphide bonds and is responsible for polymerization in many proteins (Bokhove & Jovine, 2018).

The structural integrity of the TM must be preserved throughout life to ensure proper auditory function (Richardson et al., 2008). Key components of the TM such as TECTA and TECTB are predominantly expressed during early development (Rau et al., 1999), and the TM itself undergoes progressive deterioration with age, eventually becoming less dense (Bullen et al., 2020). This age-related degradation is exacerbated in mice lacking CEACAM16—a TM protein first expressed just before the onset of hearing in mice at around postnatal day 12—highlighting the importance of ongoing molecular maintenance by specific structural components (Goodyear et al., 2019). In addition to aging, noise exposure has been shown to disrupt TM structure across multiple species, with significant morphological alterations and associated hearing loss (Adler et al., 1995; Canlon, 1987; Froymovich et al., 1995). Damage to the TM has also been documented in human temporal bone samples from individuals with histories of high noise exposure (Ishai et al., 2019). In contrast to birds, which appear to exhibit limited TM regeneration following noise (Adler et al., 1995)—most notably within basal lattice-like structures—similar regenerative capacity has not been observed in mammals.

Mutations in *TECTA* are a known cause of autosomal dominant (AD) and recessive non-syndromic hearing loss (NSHL), establishing it as a deafness gene (Verhoeven et al., 1998). Specifically, a dominant variant in *TECTA,* reported in a Spanish family and affecting the ZP domain caused mid-frequency hearing loss (Moreno-Pelayo et al., 2001), while several other variants within the same domain of TECTA are reported to cause different phenotypes, including prelingual non-progressive pan-frequency hearing loss and postlingual progressive high-frequency hearing loss (Moreno-Pelayo et al., 2001; Verhoeven et al., 1998). In studies using mouse models carrying *Tecta* variants identified in patients, mutations within the ZP domain were reported to cause hearing loss and morphological abnormalities within the TM, including disruption of highly specific structural components (Legan et al., 2014; Legan et al., 2005).

Due to reported sensitivity to sound-induced seizures in several existing *Tecta* mutant mouse models, the effects of noise trauma could not be evaluated. Specifically, three knock-in lines carrying human *TECTA* variants linked to AD hearing loss—*Tecta-C1619S/+, Tecta-C1837G,* and *Tecta-L1820F,G1824D/+*—all exhibited audiogenic seizures when exposed to octave-band white noise, even at low sound pressure levels (Legan et al., 2014), whereas wild-type littermates remained unaffected. This seizure sensitivity is likely attributable to the *S129SvEv* genetic background used in these models, as mice on this background are known to be more susceptible to audiogenic seizures than the commonly used, seizure-resistant *C57BL/6* mice. Another model, *Tecta-Y1870C* (Legan et al., 2005), was not explicitly tested for seizure sensitivity but displayed similar TM histological abnormalities, mild changes in basilar membrane tuning and distortion product otoacoustic emission (DPOAE) responses, alongside disproportionately larger deficits in neural sensitivity and neural tuning. Thus, even moderate structural defects in the TM can profoundly alter cochlear function.

Although we previously reported that *Tectb* knockout mice (*Tectb^-/-^*) display sharpened tuning, altered SSM organization and low-frequency hearing loss (Ghaffari et al., 2010; Russell et al., 2007), pathogenic variants in *TECTB* have not been previously linked to human hearing loss, leaving it the only major structural component of the TM without a confirmed deafness association, despite established roles for *CEACAM16*, *OTOA*, *OTOG*, *OTOGL*, and *TECTA* in hereditary NSHL (Booth et al., 2018; Mustapha et al., 1999; Schraders et al., 2012; Verhoeven et al., 1998; Yariz et al., 2012; Zheng et al., 2011; Zwaenepoel et al., 2002) (**Table S1**). In this study, we identified two missense variants in *TECTB*: NM_058222.3:c.674G>A, p.Cys225Tyr, hereafter abbreviated as *TECTB*-*C225Y* for the *in vivo* study, and NM_058222.3:c.853C>T, p.Arg285Cys (C225Y), in multigenerational families segregating AD mild-to-severe hearing loss. The *TECTB-C225Y* substitution affects a conserved cysteine within the ZP domain, consistent with previously reported dominant missense *TECTA* variants that disrupt ZP-domain-mediated SSM assembly and cause AD hearing loss in patients (Verhoeven et al., 1998). To study this variant, we developed a knock-in *Tectb-C225Y* mouse model and characterized auditory function and TM histology across the lifespan, and in response to noise trauma, assessing the extent and nature of the resulting deficit. Our findings establish *TECTB* as a novel human deafness gene and provide insight into the structural role of β-tectorin in TM integrity and cochlear function.

## Results

### Risk factor questionnaire for family 1

A detailed hearing loss risk factor questionnaire was completed by all available family members prior to genetic analysis. Affected status was determined by pure-tone audiometry (**Table S2**). Although none of the affected individuals reported excessive noise exposure, accurate assessments of noise exposure are difficult and may be underestimated in rural agricultural settings. None of the individuals who completed the questionnaire reported a history of maternal infection, otorrhea, alcohol consumption during pregnancy or exposure to aminoglycoside antibiotics such as gentamicin. Half of the respondents reported exposure to pesticides, including four of six (66.7%) affected individuals (II:5, III:2, III:5, and III:6), although the timing and extent of exposure were not specified. None of the six affected individuals reported birth defects or prematurity. Four of six (66.7%) affected individuals (II:4, II:5, III:3, and III:6) reported balance difficulties, although these were not formally evaluated with videonystagmography or other clinical vestibular testing. One individual (II:5) reported a history of otitis media that resolved with treatment, and another (II:4) reported a temporary threshold shift following noise exposure (16.7% of affected individuals for each observation).

### Pure-tone audiometry from family 1 shows stable, mild to severe, sensorineural hearing loss

The male proband from family 1 (II:4) was born to healthy, non-consanguineous parents (**Figure 1a**) and was in his late 60s at the time of his initial evaluation for this study. Three of the proband’s five children have hearing loss, as well as his sibling and other distant family members. Newborn hearing screening was not performed. Hearing loss was reported to start between 6 and 15 years of age in all affected individuals. The proband and two of the affected family members underwent a follow-up evaluation more than a decade after enrollment in the study, providing an opportunity to assess hearing loss progression.

**Figure 1.**
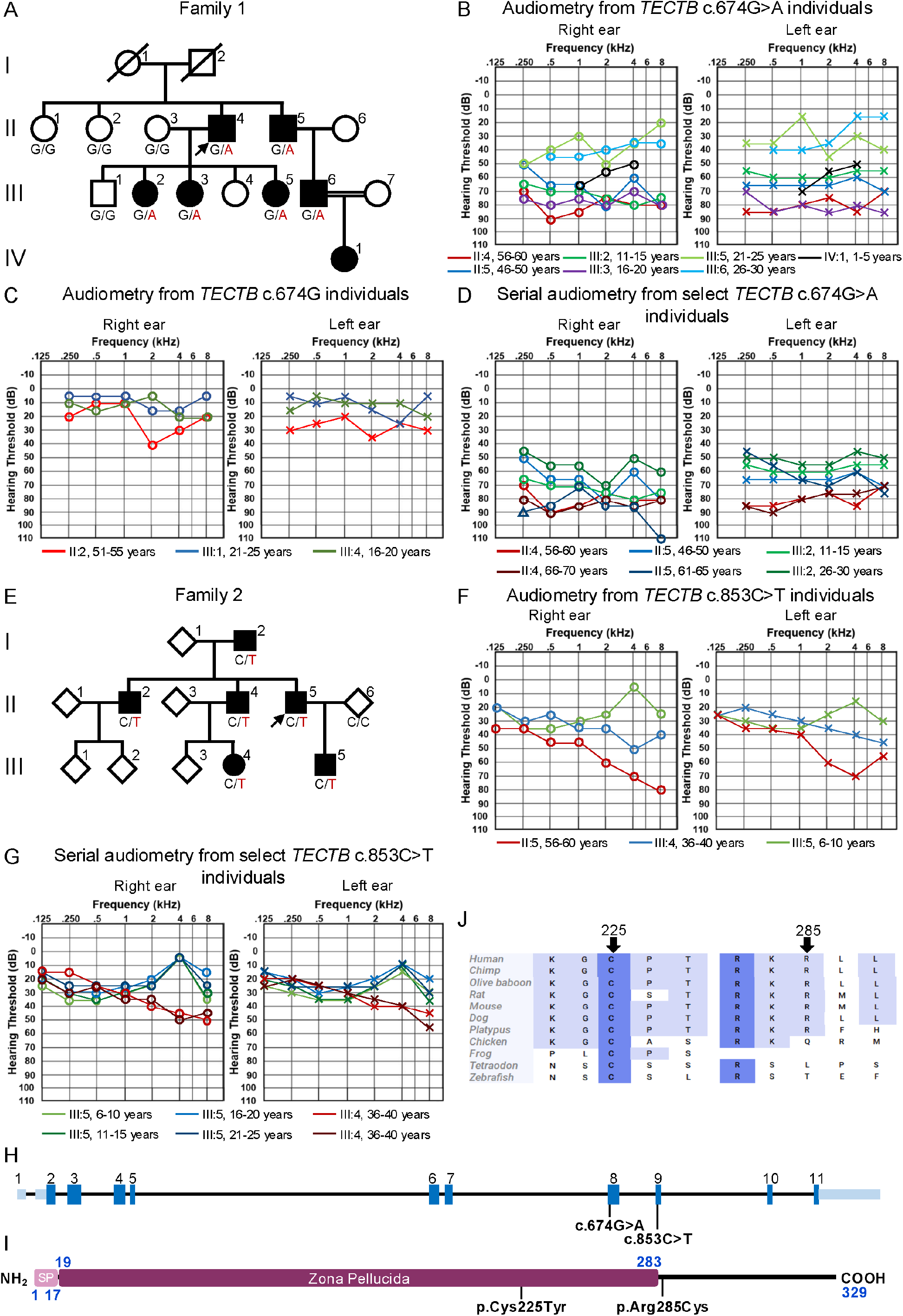
Pedigree, audiological data and TECTB variant schematic on gene and protein levels. (**a**) An abbreviated pedigree of the multigenerational Family 1 showing segregation of the heterozygous *TECTB* c.674G>A, p.Cys225Tyr (C225Y) variant. Filled symbols indicate affected individuals; open symbols indicate unaffected individuals. The proband II:4 is denoted with an arrow. Deceased individuals are indicated by a diagonal line. The variant nucleotide substitution “A” is shown in red. (**b**) Pure-tone audiograms of affected individuals II:4 (*red*), II:5 (*dark blue*), III:2 (*dark green*), III:3 (*purple*), III:5 (*light green*), III:6 (*light blue*), and IV:1 (*black*), with corresponding age ranges at the time of testing in the legend below. Air-conduction thresholds in dB hearing level are represented by circles (*right ear*) and crosses (*left ear*). (**c**) Pure-tone audiograms of unaffected individuals II:2 (red), III:1 (blue), and III:4 (green) carrying the reference allele, with ages at the time of evaluation presented in the legend below. Symbols as in panel *b*. (**d**) Follow-up pure-tone audiograms from II:4, II:5 and III:2 that span 12, 16, and 16 years, respectively. (**e**) The pedigree of Family 2 showing segregation of the heterozygous *TECTB* c.853C>T, p.Arg285Cys (R285C) variant. The proband II:5 is denoted with an arrow. Diamonds represent an undisclosed sex, otherwise, symbols are as described in panel *a*. (**f**) Pure-tone audiograms of affected individuals II:5 (red), II:4 (blue), and III:5 (green), with corresponding age ranges at the time of testing in the legend below. Symbols as in panel *b*. (**g**) Follow-up pure-tone audiograms from III:5 and III:4 spanning 13 (III:5) and 1 year (III:4), respectively. (**h**) Schematic representation of the *TECTB* (NM_058222.3) variants, with coding exons shown in dark blue boxes and untranslated sequences in light blue. The identified variants map to exons 8 and 9. (**i**) Domain structure of the TECTB protein, showing the signal peptide (SP, *light maroon*), and the conserved zona pellucida domain (ZP, *maroon*). The location of the C225Y and R285C substitutions are indicated with black bars. (**j**) Multiple sequence alignment of TECTB orthologues highlighting the conserved residues at positions 225 and 285 (*arrows*), substituted in the families.

Pure-tone audiometry of participating individuals revealed symmetrical, bilateral sensorineural hearing loss (SNHL) (**Figure 1b**). Follow up measurements are available from II:4, II:5 and III:2 that span 12, 16, and 16 years, respectively (**Figure 1d**). At their initial assessment at age 56-60, the proband (II:4) showed severe SNHL (PTA0.5-4kHz 81.25 dB HL), which had remained unchanged at the age of 66-70 (PTA0.5-4kHz 80 dB HL). One of his children also demonstrated severe hearing loss (III:2, PTA0.5-4kHz 73.75 dB HL) at age 11-15, which measured as moderate hearing loss at age 26-30 (PTA0.5-4kHz 51.25 dB HL). Another child (III:5, PTA0.5-4kHz 31.25 dB HL) at the age of 21-25 years and a cousin (III:6, PTA0.5-4kHz 32.50 dB HL) at the age of 26-30 presented with mild hearing loss. Moderate hearing loss was observed in another child (III:3, PTA0.5-4kHz 58.75 dB HL) at age 16-20, a distant relative (IV:1, PTA1.0-4kHz 58.33 dB HL) at age 1-5, and the proband’s sibling (II:5, PTA0.5-4kHz 63.75 dB HL) at age 46-50, which had remained unchanged at the age of 66-70 (PTA0.5-4kHz 62.5 dB HL). Serial audiometry was not available for a subset of individuals but collectively, the available PTA0.5-4kHz data reveal substantial intrafamilial variability, ranging from mild to severe hearing loss, without progression. Otoacoustic emissions were bilaterally absent in the most recent measurements from II:4, II:5 and III:2 at the age of 66-70, 61-65, and 26-30 years, respectively.

### Pure-tone audiometry from family 2 shows mild to moderate SNHL

The male proband from family 2 (II:5) was born to a hearing-impaired parent, consistent with AD inheritance (**Figure 1e**). The proband reported onset of hearing loss at approximately 6-10 years of age with gradual progression over time. Pure-tone audiometry at 56-60 years of age demonstrated bilateral, symmetrical, moderate SNHL with a sloping audiogram configuration (PTA0.5-4kHz 45 dB HL) (**Figure 1f**). An affected child (III:5) showed mild hearing loss with relatively flat audiogram and preserved hearing thresholds at 4 kHz (PTA0.5-4kHz 23.75 dB HL) at 6-10 years of age. Hearing remained stable at the most recent assessment at 21-25 years of age (PTA0.5-4kHz 20dB HL) (**Figure 1g)**. DPOAEs and transient otoacoustic emissions were present when measured at 16 years of age. Another affected family member (III:4) demonstrated mild bilateral hearing loss with gently sloping audiograms (PTA0.5-4kHz 32.25 dB HL) at 36-40 years of age **(Figure 1f**), which remained stable one year later (PTA0.5-4kHz 32.5 dB HL) **(Figure 1g**). Collectively, the available audiometric data indicate mild-to-moderate SNHL with intrafamilial variability.

### The *TECTB-C225Y* variant segregates with AD NSHL in family 1

Gene panel sequencing and analysis of 74 hearing loss-associated genes was performed on genomic DNA from the proband (family 1, II:4) but yielded no diagnostic findings, leaving the family genetically undiagnosed. To further investigate the possible genetic cause, two-point linkage analysis of the genome scan markers was conducted, revealing several loci with elevated logarithm of the odds (LOD) scores. A genome-wide multipoint LOD score representation of 22 autosomes identified five peaks above 2.0 on chromosomes 2, 9, 10, 14 and 15 (**Figure S1a**), with a maximum multipoint LOD score of 2.0569 at rs7098615 and rs17129748 on chromosome 10 (**Figure S1b**) – a region that includes *TECTB* and two other genes (*GPAM*, and *ACSL5*) (**Figure S1c**).

Subsequent exome sequencing of the genomic DNA of the proband generated 195,746,072 total reads, of which 158,815,063 aligned with high quality, and 85.81% achieved a target coverage of 20x. Analysis of linkage intervals, as well as exome-wide analysis identified a heterozygous *TECTB* NM_058222.3:c.674G>A, p.Cys225Tyr missense variant in exon 8 (**Figure 1h**, **Table 1**, **Figure S2**). This variant results in an amino acid substitution of a small, polar cysteine to a larger aromatic tyrosine in the ZP domain (**Figure 1i**), a region known to mediate extracellular matrix assembly. Multiple *in silico* prediction tools unanimously classified the variant as deleterious/damaging (**Table 1**), and phyloP analysis confirmed conservation of the affected nucleotide across multiple species (**Figure 1j**). The *TECTB* c.674G>A, p.Cys225Tyr variant was absent from major population databases, including *gnomAD* v4.1.0 and the *All of Us* Data Browser (All of Us Research Program Genomics, 2024).

**Table 1.** Summary of *TECTB* variants and analysis results.

|  | Family 1 | Family 2 |
| --- | --- | --- |
| Molecular genetics summary |  |  |
| Chr10 g. position (GRCh37) | g.114,057,829G>A | g.114,059,268C>T |
| Chr10 g. position (GRCh38) | g.112,298,071G>A | g.112,299,510C>T |
| *TECTB* c. position (NM_058222.3) | c.674G>A | c.853C>T |
| *TECTB* p. position (NP_478129.1) | p.Cys225Tyr | p.Arg285Cys |
| Zygosity | Heterozygous | Heterozygous |
| *In silico* predictions |  |  |
| phyloP [-19.0;11.0] | 7.93 | 4.09 |
| SIFT | Deleterious | Deleterious |
| PolyPhen-2 | Probably Damaging | Probably Damaging |
| FATHMM | Damaging | Damaging |
| MutationTaster | Deleterious | Deleterious |
| REVEL | Damaging | Damaging |
| CADD | 28.9 | 27.7 |
| ClinPred | Damaging | Damaging |
| Splicing Predictions | NR | NR |
| gnomAD v4.1.0 MAF | 0 | 1.2e-6 |
| All of Us Data Browser | 0 | 0 |

Abbreviations: MAF, minor allele frequency; NR, Not remarkable

Next, we performed a segregation analysis using Sanger sequencing revealing complete co-segregation of the *TECTB* c.674G>A, p.Cys225Tyr variant with the hearing loss phenotype across the family (**Figure 1a**). Twelve additional rare variants identified in the exome sequencing data were excluded based on lack of segregation or predicted pathogenicity (**Table S3**).

Among the three unaffected individuals carrying only the reference allele – II:2 (age 51-55), III:1 (age 21-25) and III:4 (age 16-20) – PTA0.5-4kHz were within normal limits (**Figure 1c**). Of note, individual II:2 showed elevated thresholds at 2 kHz (40 and 35 dB in right and left ears, respectively), though these findings fall within the expected range of age-related hearing decline and are not considered disease-associated.

### The *TECTB-R285C* variant segregates with AD NSHL in family 2

Initial sequencing using the *TruSight One* panel on genomic DNA from the proband (II:5), affected child (III:5) and unaffected partner (II:6) did not identify suspect variants in established hearing loss-associated genes. Re-analysis identified a heterozygous *TECTB* c.853C>T, p.Arg285Cys missense variant in the proband and his affected child. To further investigate the genetic basis of hearing loss in the family, exome sequencing was subsequently performed on the proband (II:5), his affected sibling (II:2) and affected child (III:5). Exome filtering and segregation analysis confirmed the presence of the *TECTB* c.853C>T, p.Arg285Cys variant in all affected individuals (**Figure 1e**). PhyloP analysis showed strong evolutionary conservation (**Figure 1j**). The variant was either absent or extremely rare in population databases, including gnomAD v4.1.0 and *All of Us* Data Browser. Sanger sequencing demonstrated complete segregation in six affected individuals. The identification of a second independently segregating *TECTB* missense variant in a separate multigenerational family further supports *TECTB* as a cause of AD NSHL.

### *Tectb^C225Y/C225Y^* mice show a severe hearing deficit

The *Tectb-C225Y* mouse was created using the CRISPR-Cas9-mediated genome editing approach, introducing a c.674G>A (TGC to TAC) substitution in exon 8 of the *Tectb* gene to mimic the human C225Y variant (**Figure S3**). The initial editing was performed in *C57BL6N* background, which carries the *Cdh23^Ahl^*allele known to cause early-onset age-related hearing loss. Following validation, the mutation was backcrossed onto a *C57BL6-Cdh23^Ahl+^* mouse strain (JAX#002756) with the corrected *Cdh23* allele to preserve normal hearing. The resulting TECTB-C225Y mice appeared healthy, exhibited normal fertility, and produced offspring consistent with Mendelian ratios.

To evaluate the functional impact of the *TECTB-C225Y* substitution on cochlear function, *Tectb^C225Y/C225Y^*, *Tectb^C225Y/+^*, and wild-type mice were assessed using auditory brainstem response (ABR, **Figure 2a, and S4**) and DPOAE (**Figure 2b**) testing at 5 weeks and at 3, 6, 9, 12, and 18 months of age. Across the assessed time points, *Tectb^C225Y/C225Y^* mice exhibited severe, progressive SNHL, with average ABR threshold of approximately 60 dB SPL at 11.2 kHz (best frequency) at 5 weeks, deteriorating to profound deafness in most animals by 12 months. In contrast, *Tectb^C225Y/+^* mice retained normal ABR thresholds across all time points and frequencies, indistinguishable from wild-type and consistent with normal age-related decline. Average ABR waveforms across all tested frequencies and time points for all three genotypes are presented in **Figure S4**, providing a comprehensive overview of the progressive changes in response morphology. Of note, *Tectb^C225Y/+^*and wild-type waveforms closely align throughout the matrix; to confirm this quantitatively, we measured peak I latency and amplitude in response to 11.2 kHz pure tone stimulation at 80 dB SPL in 12-month-old mice — the frequency and time point where the average traces most suggested a potential amplitude difference — and found no statistically significant difference between genotypes (**Figure S5**). Average waveforms from 18-month-old mice revealed a lower wave I peak amplitude in *Tectb^C225Y/+^* mice compared to wild-type at this age (**Figure S4**), however, individual waveforms at this age were too noisy and greatly reduced in amplitude to allow for a reliable quantitative analysis.

**Figure 2.**
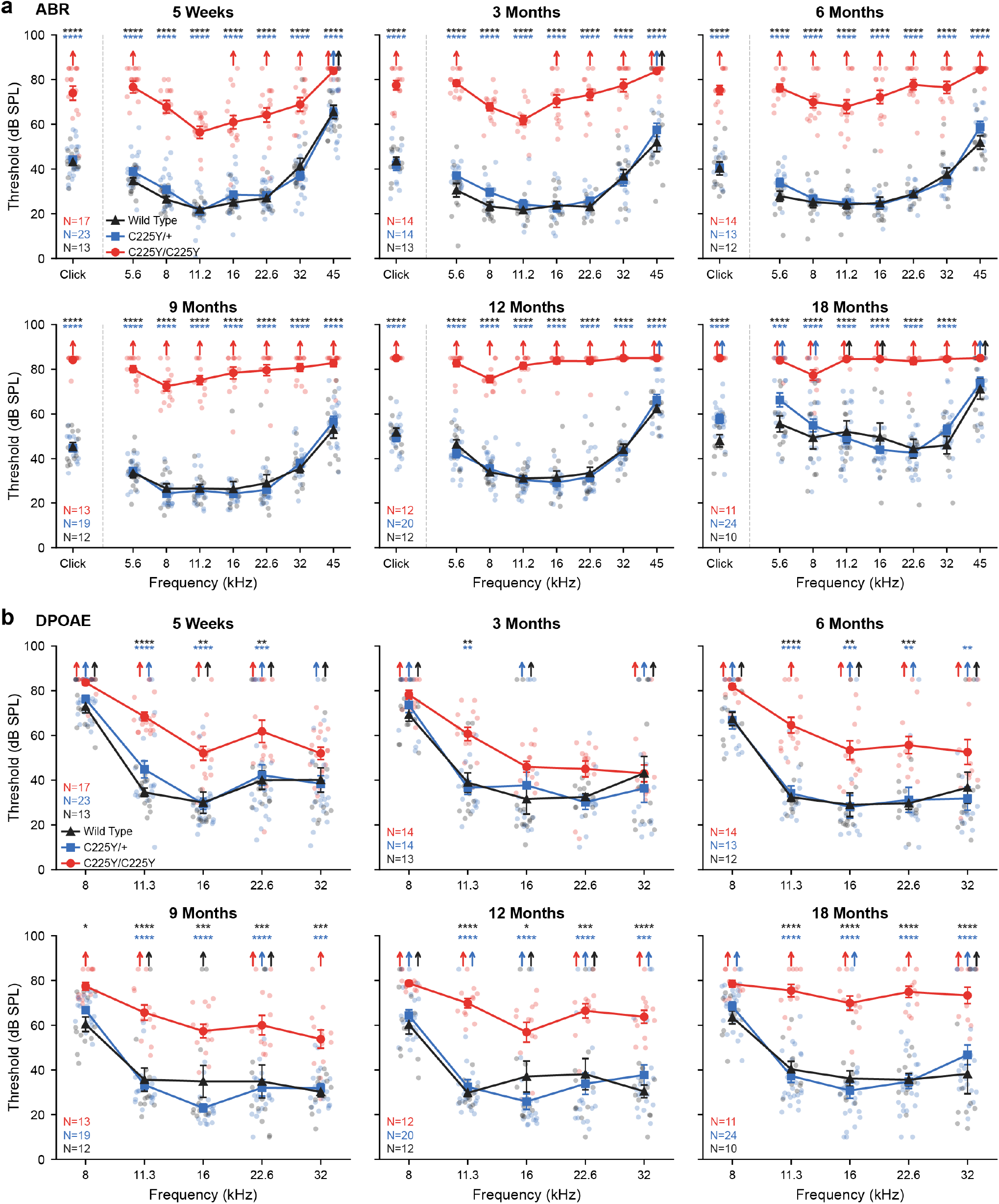
*Tectb^C225Y/C225Y^* mice exhibit a severe hearing deficit. (**a**) Auditory brainstem response (ABR) and (**b**) distortion product otoacoustic emission (DPOAE) thresholds of *Tectb-C225Y* mice tested at 5 weeks, and 3, 6, 9, 12, and 18 months of age. Data are shown as individual values with an overlaid mean ± SEM. Upward-pointing arrows indicate that at the highest sound pressure level tested (80 dB SPL), at least one mouse in the group had no detectable response at that frequency; these values were entered as 85 dB for analysis. *Tectb^C225Y/C225Y^* mice exhibited significantly higher ABR and DPOAE thresholds than both *Tectb^C225Y/+^* (*blue asterisks*) and wild type (*black asterisks*) mice across all tested ages. No significant differences were observed between *Tectb^C225Y/+^*and wild-type mice. The number of animals per group is indicated within each plot. *Statistical analysis*: two-way ANOVA with a matched design between frequencies for each mouse. Šidák’s multiple comparison test was to evaluate differences between genotypes at each frequency; *ns* – not significant, * p<0.05, ** p<0.01, *** p<0.001, **** p<0.0001.

Somewhat surprisingly, DPOAE threshold elevations in *Tectb^C225Y/C225Y^* mice were substantially less severe than the corresponding ABR threshold elevations, with partial preservation at younger ages and lower frequencies despite the severe ABR phenotype, suggesting residual OHC function. Nonetheless, DPOAE thresholds progressively increased with age, particularly at higher frequencies. In contrast, *Tectb^C225Y/+^* mice showed DPOAE thresholds nearly indistinguishable from wild-type controls at all tested ages, further underscoring the subtlety of the baseline phenotype in heterozygous animals.

To further characterize the divergence between ABR and DPOAE responses in *Tectb^C225Y/C225Y^* mice, we examined ABR waveforms and suprathreshold growth functions at 5 weeks of age — the earliest time point at which both measures remained partially preserved in homozygous mice, providing sufficient dynamic range for meaningful comparison (Figure 3). ABR waveforms evoked at 80 dB SPL revealed markedly reduced wave I amplitudes across all tested frequencies in *Tectb^C225Y/C225Y^* mice, with a possible shift in peak latency (Figure 3a). Given the reduced wave I amplitudes in *Tectb^C225Y/C225Y^* mice, we considered whether the summating potential (SP) — a DC receptor potential reflecting predominantly IHC input when recorded with alternating-polarity stimulation (Pappa et al., 2019; Zheng et al., 1997), which precedes wave I in the ABR waveform — might be disproportionately affected, potentially implicating IHC dysfunction. To assess this, we scaled average traces to matching peak I amplitudes, allowing direct comparison of relative SP magnitude across genotypes (Figure 3a**, insets**). This normalization revealed no apparent difference in SP proportion between genotypes, suggesting that IHC transduction itself is not selectively impaired by the TECTB-C225Y variant.

**Figure 3.**
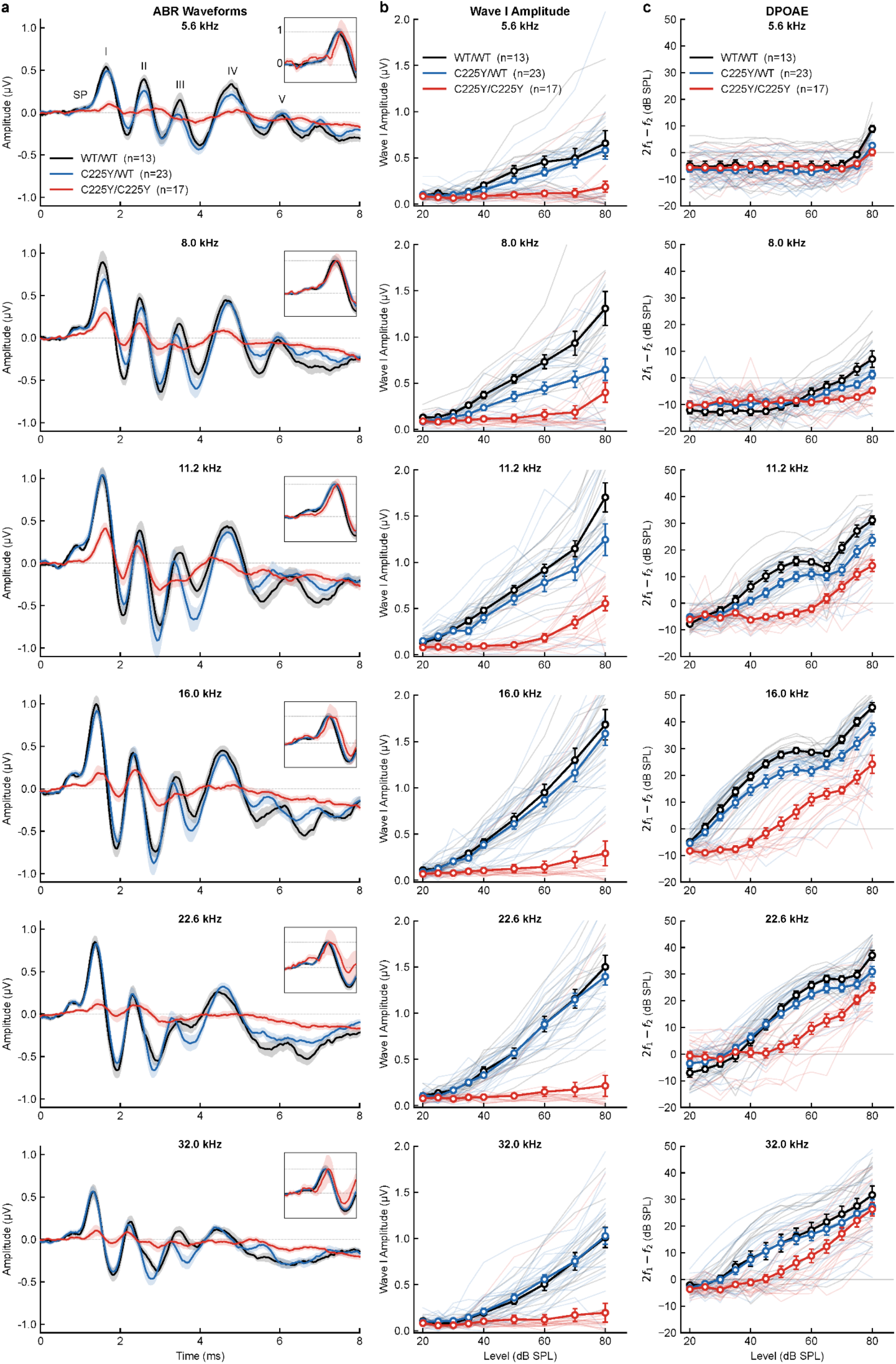
ABR deficits in *Tectb^C225Y/C225Y^* mice exceed DPOAE deficits, particularly at high frequencies. (a) Average ABR from *Tectb-C225Y* mice tested at 5 weeks of age in response to 5.6, 8.0, 11.2, 16, 22.6, and 32 kHz at 80 dB SPL. *Tectb^C225Y/C225Y^* mice show markedly reduced wave I amplitudes across all frequencies. Shaded areas represent SEM. Insert shows all average traces scaled to matching peak 1 amplitudes, allowing comparison of relative summating potential (SP) amplitude. No clear difference in SP proportion to wave I was observed between genotypes. (**b**) and (**c**), Average growth functions for ABR wave I amplitude (*b*) and DPOAE (*c*) from 5-week-old cohort at 5.6, 8.0, 11.2, 16, 22.6, and 32 kHz. The deficit in wave I amplitude in *Tectb^C225Y/C225Y^* mice was greater than the deficit in DPOAE amplitude, with the largest mismatch at higher frequencies. Individual growth functions shown as lighter lines alongside averages.

When ABR wave I amplitude growth functions were compared directly with DPOAE growth functions at the same frequencies, the deficit in *Tectb^C225Y/C225Y^*mice was consistently larger for ABR than for DPOAE, with the greatest mismatch at higher frequencies (Figure 3b**,c**). This pattern is consistent with a largely preserved OHC function and OHC bundle stimulation, suggesting that mechanical stimulation of IHC stereocilia is disproportionately impaired. Consistent with this interpretation, qualitative assessment of IHC afferent synapse counts at 10 weeks of age showed no overt difference between genotypes, indicating that the ABR deficit in *Tectb^C225Y/C225Y^* mice is unlikely to result from synaptopathy (**Figure S6**).

Overall, these findings contrast with the autosomal-dominant inheritance observed in both families, segregating the *TECTB-C225Y* and *TECTB-R285C* alleles, in which a single copy of the variant allele is sufficient to cause significant levels of hearing loss. To explore the underlying cause of this discrepancy and to evaluate the validity of the mouse model in recapitulating the human phenotype, we next performed histological analysis of the TM in mice.

### TECTB-C225Y increases TM volume and decreases matrix-associated staining

Following ABR and DPOAE testing, cochleae were fixed, resin-embedded and sectioned to allow morphological analysis of the TM. A previously established sectioning approach (Cheatham et al., 2014; Goodyear et al., 2019) using 1 µm sections stained with toluidine blue was used to visualize TM profiles corresponding to the 4, 8, 20 and 40 kHz frequency regions of the cochlea on a single mid-modiolar cross-section (Figure 4a).

**Figure 4.**
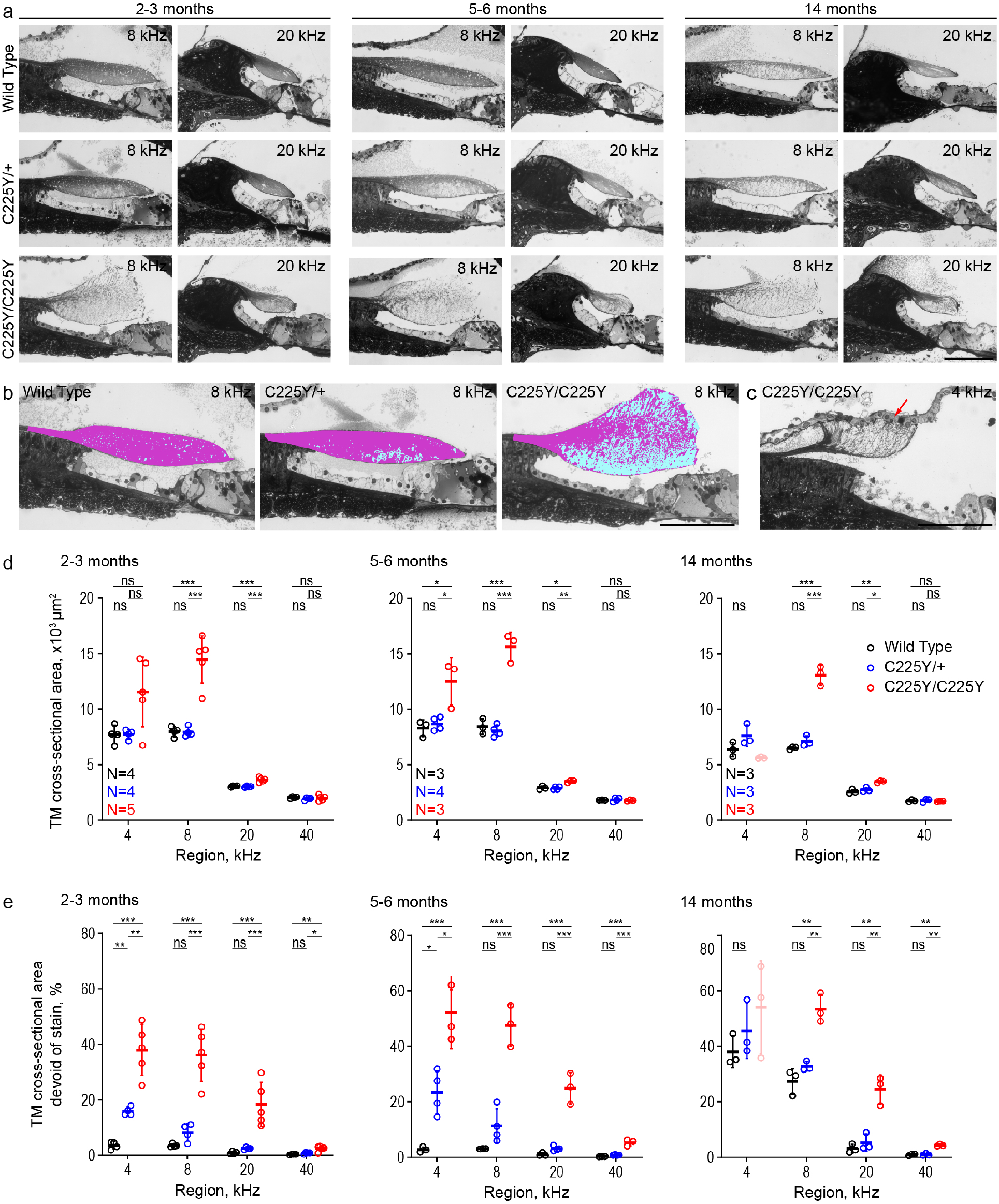
TM morphology is disrupted in mice expressing TECTB-C225Y. (**a**) Toluidine blue-stained 1 µm resin sections from the 8 and 20 kHz cochlear regions of wild-type, *Tectb^C225Y/+^* and *Tectb^C225Y/C225Y^* mice at 2-3 months of age. (**b**) Representative images showing the adaptive thresholding method used to quantify TM cross-sectional area and staining density. Images correspond to the 8 kHz region from 2–3-month-old mice shown in *A*. *Cyan*, unstained portion of TM. *Magenta*, stained portion of TM. (**c**) Example of a cochlear cross-section from the 4 kHz region of a 14-month-old *Tectb^C225Y/C225Y^* mouse, illustrating fusion of the disrupted TM with Reissner’s membrane (*red arrow*). *Scale bars,* 100 µm. (**d**) Quantification of TM cross-sectional area across genotypes and regions. (**e**) Percentage of the TM cross-sectional area that is unstained. Values for *Tectb^C225Y/C225Y^* TMs at 4 kHz (panels *d-e*), indicated in pink, were aberrated by attachment of the TM to Reissner’s membrane. The inclusion of these values did not affect the statistical significance levels calculated. *Statistical analysis:* One-way ANOVA was performed for each frequency, followed by Šidák’s multiple comparisons test between genotypes with Benjamini-Hochberg correction applied within each age group. Percentage of unstained area values were arcsine square root-transformed prior to statistical analysis. ns – not significant, * p<0.05, ** p<0.01, *** p<0.001.

At two months of age, *Tectb^C225Y/C225Y^* cochleae exhibited, as expected, a gradient in TM size and shape from apex to base; however, severe structural abnormalities were already evident. The cross-sectional area of the TM was moderately enlarged in the 20 kHz region and was considerably increased in the 8 kHz region across all time points. Despite this enlargement, toluidine blue staining was more diffusely distributed in *Tectb^C225Y/C225Y^* samples, suggesting the change in size reflected an increase in areas devoid of staining rather than an increase in TM material (Figure 4a).

By contrast, the TM morphology in *Tectb^C225Y/+^* cochleae was grossly similar to that of wild-type controls. However, an increase in regions devoid of toluidine blue staining was consistently observed in the central body of the TM, particularly in the apical, low-frequency region of the cochlea. The lower matrix density observed in regions of the TM in young *Tectb^C225Y/+^* mice resembles that seen to a greater extent in older mice by 14 months of age. Whether this is a consequence of early-onset degradation or failure in matrix formation or deposition remains to be resolved (Figure 4a).

To quantitatively assess TM morphology, we performed blinded morphometric analysis using adaptive thresholding, measuring cross-sectional area and staining intensity, with the latter used as a measure of matrix content (Figure 4b). For each image, the outer border of each TM was manually traced to calculate the area (Figure 4d). A threshold was then applied to mask the stained regions of the TM, allowing quantification of unstained area (see Figure 4b compared to Figure 4a images of 2-3-month-old TMs at 8 kHz), expressed as a percentage of the total TM cross-sectional area (Figure 4e). This metric served as a proxy measure of matrix content within the TM.

At the apical end of *Tectb^C225Y/C225Y^* cochleae, the TM was frequently found to be completely disintegrated or displaced and bound to Reissner’s membrane (Figure 4c *red arrow*), precluding reliable quantification in this region. Therefore, measurements from this location of *Tectb^C225Y/C225Y^* cochleae were excluded from further analysis. Quantification of all other images revealed a significant increase in TM cross-sectional area in *Tectb^C225Y/C225Y^* mice relative to wild-type mice across all cochlear locations except the extreme basal, 40 kHz region (Figure 4d). *Tectb^C225Y/+^* mice, by contrast, showed no significant difference in TM area compared to wild-type mice.

In contrast to area measurements, the proportion of the TM cross-section lacking stain—an indirect measure of TM matrix content—was elevated in *Tectb^C225Y/C225Y^* and to a lesser extent in *Tectb^C225Y/+^* mice as compared to wild-type controls (Figure 4e). Notably, this increase was especially prominent in the apical, low-frequency regions of the cochlea, even in *Tectb^C225Y/+^*mice that otherwise displayed normal TM area values. An age-associated increase in cross-sectional area devoid of stain was also observed in wild-type mice at later time points, suggesting that the TECTB-C225Y variant might accelerate structural degradation of the TM (Figure 4e).

### TECTB-C225Y disrupts Hensen’s stripe, the marginal band, and the SSM

In addition to the observed changes in TM thickness and stain density, the TECTB-C225Y variant also caused subtle but consistent disruptions in structural and ultrastructural features of the TM (Figure 5). Specifically, abnormalities were noted in the morphology of the marginal band and Hensen’s stripe (Figure 5a, *arrows* and *arrowheads,* respectively)—two key features of the TM—in both *Tectb^C225Y/+^* and *Tectb^C225Y/C225Y^* mice. These differences were most pronounced in the basal, high-frequency region of the TM, where such structural features are most readily identified in cross-sections.

**Figure 5.**
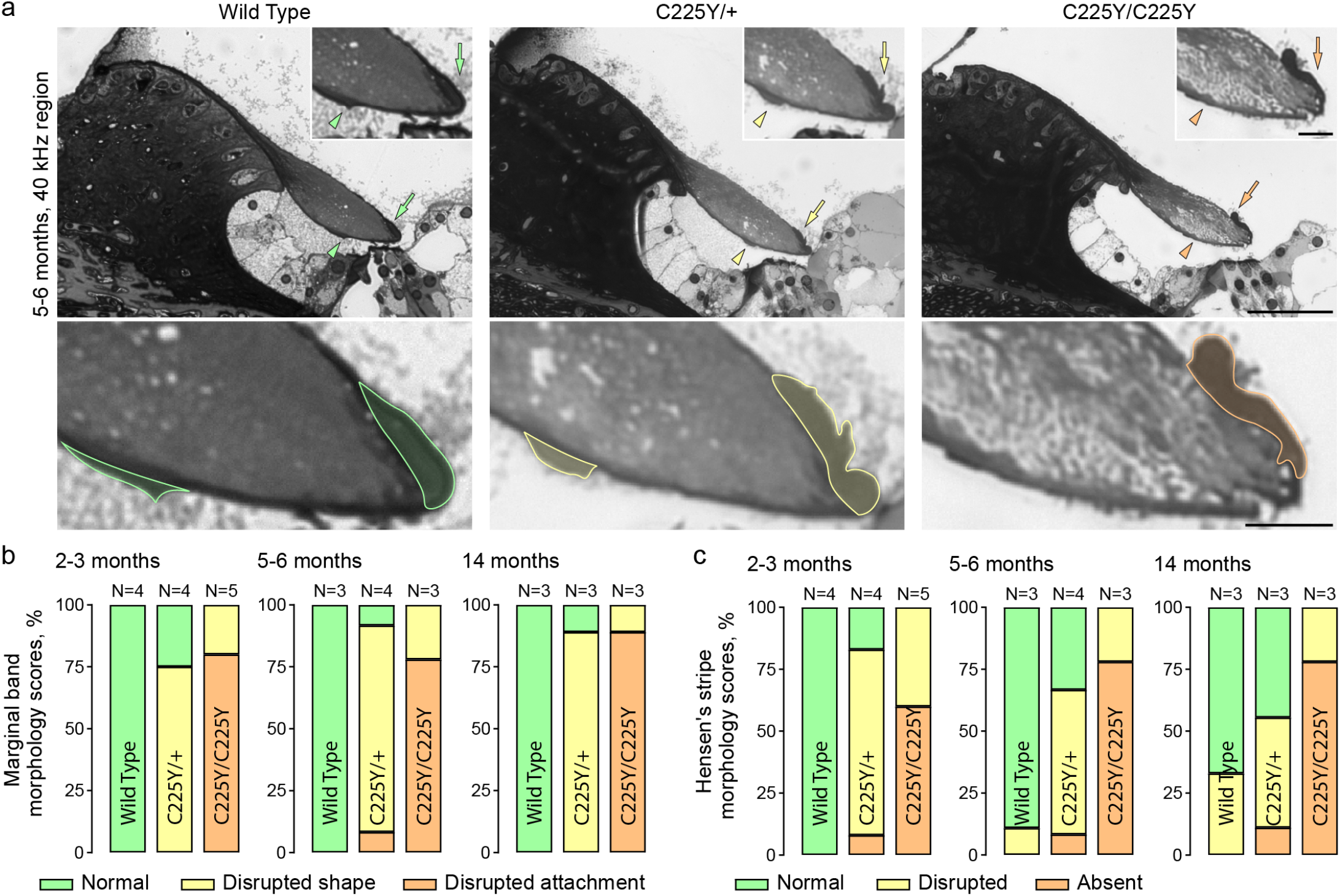
TECTB-C225Y disrupts TM ultrastructure. (**a**) Representative micrographs of toluidine blue-stained 1 µm resin sections from the 40 kHz region of 5-6-months-old wild-type, *Tectb^C225Y/+^* and *Tectb^C225Y/C225Y^* cochleae. Arrowheads indicate Hensen’s stripe, arrows indicate the marginal band. Both structures are outlined in lower panel for clarity. *Scale bars,* 50 µm (*top*) and 10 µm (*insets and bottom*). (**b**) and (**c**), pooled morphology scores from blinded reviewers assessing the integrity of the marginal band (*b*) and Hensen’s stripe (*c*) in TM cross-sections from the 40 kHz region. Scoring was based on structural continuity and deviation from typical shape. The number of animals per group is indicated within each plot.

To quantify these observations, three investigators independently evaluated marginal band and Hensen’s stripe morphology in cross-sections at the 40 kHz region while blinded to genotype. The pooled scores are presented in Figure 5b-c. While individual cross-sections showed some variability, morphology scores consistently segregated by genotype, suggesting a genotype-dependent structural deficit.

In *Tectb^C225Y/+^* mice, the TM marginal band frequently appeared less smooth and less rounded compared to the uniform, curved morphology observed in wild-type mice (Figure 5a, *arrows*). These deficits were further exaggerated in *Tectb^C225Y/C225Y^* mice, where the marginal band was often raised or deformed at its upper edge. Graded disruption was also evident in Hensen’s stripe, ranging from altered morphology in *Tectb^C225Y/+^*mice to complete absence in some *Tectb^C225Y/C225Y^* sections (Figure 5a, *arrowheads*). Notably, occasional disruption of Hensen’s stripe morphology was also observed in older wild-type mice (Figure 5c), further suggesting a potential age-related degradation component, or that these phenotypes could be indicative of accelerated aging of the TM in *Tectb^C225Y/+^* mice.

Finally, the TM cross-sections were evaluated using transmission electron microscopy (TEM). Ultrastructural analysis revealed that *Tectb^C225Y/C225Y^*mice lacked the stereotypical SSM architecture characteristic of wild-type and *Tectb^C225Y/+^* TMs, indicating a profound disruption of matrix organization (Figure 6). Interestingly, anti-TECTB immunolabeling in cochlear cryosections demonstrated that TECTB-C225Y protein is still present in the TM of *Tectb^C225Y/C225Y^* mice (**Figure S7**), despite the complete absence of the SSM, suggesting that the C225Y substitution does not prevent the secretion of TECTB or its localization to the TM, but likely disrupts an ability to interact with other proteins thereby preventing assembly of the SSM. Immunolabeling was also used to assess whether the TECTB-C225Y protein influences the presence and distribution of three other components of the TM, TECTA, CEACAM16 and COL9A, but no obvious differences were detected in either the *Tectb^C225Y/+^* or the *Tectb^C225Y/C225Y^*mice relative to that in wild-type *Tectb^+/+^* mice at six weeks of age (**Figure S8**). The absence of SSM in *Tectb^C225Y/C225Y^*mice is consistent with the profound ABR and DPOAE deficits. In contrast, *Tectb^C225Y/+^*mice showed only mild morphological disruptions in the TM, with no apparent deficits in ABRs and DPOAEs. The mild structural phenotype accompanied by normal auditory thresholds in *Tectb^C225Y/+^* mice prompted us to test whether environmental stress factors, such as noise exposure, might unmask latent auditory vulnerability.

**Figure 6.**
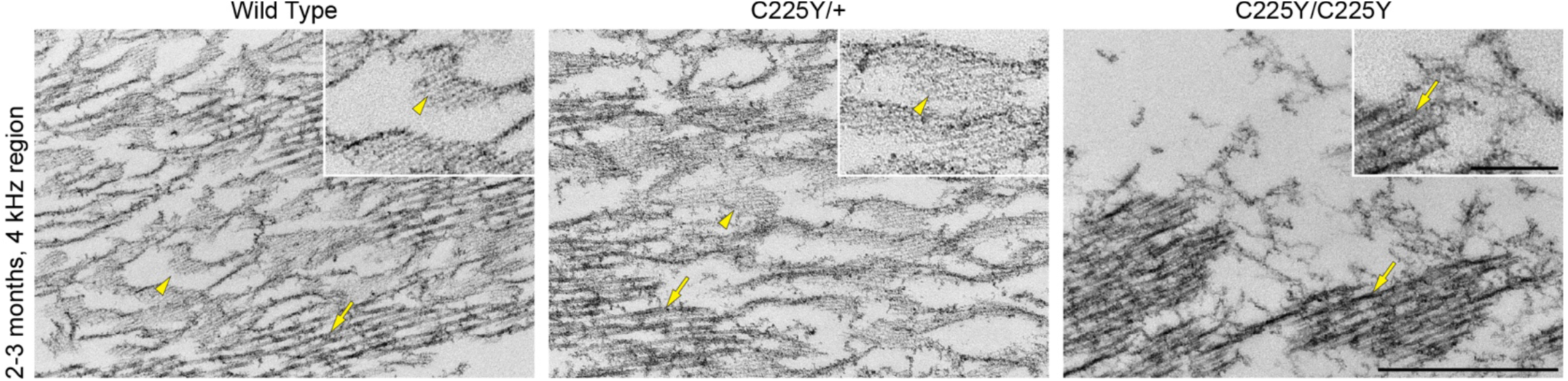
*Tectb^C225Y/C225Y^* tectorial membranes lack striated-sheet matrix. Transmission electron micrographs from the central body of tectorial membranes in the 4 kHz region of 2–3-month-old mice. *Arrows* point to collagen fibers; *arrowheads* point to striated-sheet matrix. *Scale bars*, 400 nm (*main panels*) and 100 nm (*insets*).

### *Tectb^C225Y/+^* mice exhibit increased susceptibility to noise insult

Family 1 exhibited a clear autosomal-dominant phenotype. In contrast, *Tectb^C225Y/+^* mice raised in a relatively quiet, controlled animal facility environment displayed normal ABR and DPOAE thresholds despite mild morphological defects in the TM. We reasoned that, unlike laboratory mice, humans are routinely exposed to environmental noise, a condition that might unmask a latent vulnerability conferred by the *TECTB-C225Y* variant, potentially explaining the apparent species discrepancy.

To test this hypothesis, we exposed cohorts of 16-week-old and 18-month-old mice to 8-16 kHz octave-band noise at 100 dB SPL for 2 hours using a previously established exposure paradigm that reliably produces temporary threshold shifts (TTS) in wild-type mice of this age (Jensen et al., 2015; Kujawa & Liberman, 2006). ABRs and DPOAEs were measured before exposure and again at 2 days and 2 weeks post-exposure (Figures 7 **and 8**). At both ages, and at both post-exposure time points, *Tectb^C225Y/+^*mice exhibited significantly greater ABR threshold shifts than wild-type littermates. While 16-week-old wild-type mice exhibited near-complete recovery by two weeks—a pattern consistent with TTS—*Tectb^C225Y/+^*mice failed to recover, instead developing a permanent threshold shift (PTS) of up to 28.46±5.13 dB SPL. In the 18-month-old cohort, both wild-type and *Tectb^C225Y/+^* mice exhibited a PTS, though the threshold shifts were significantly larger in the *Tectb^C225Y/+^*group. ABR waveforms recorded before and after noise exposure showed more substantial wave I amplitude reduction in *Tectb^C225Y/+^* mice at post-noise time points across tested frequencies, with an apparent increase in peak latency (Figure 7c**, 8c**). We next plotted the ABR wave I growth function in 16-week-old mice before and 2 weeks post-noise trauma, revealing slightly reduced wave I amplitude growth functions in wild-type mice, with a more severe deficit observed in *Tectb^C225Y/+^* mice at high frequencies (Figure 7d). To assess pre- to post-noise amplitude changes, wave I amplitudes were averaged across the 40–80 dB SPL stimulus range for 16, 22.6 and 32kHz and presented as bar graphs for each genotype (Figure 7b). Although individual ABR waveforms in 18-month-old mice were too noisy and greatly reduced in amplitude to allow the wave I growth function analysis performed in the 16-week cohort, average waveforms revealed a greater reduction in wave I amplitude in *Tectb^C225Y/+^* mice two weeks after noise exposure (Figure 8c**,d**, 22.6-32kHz), consistent with the findings reported for the 16-week-old cohort.

**Figure 7.**
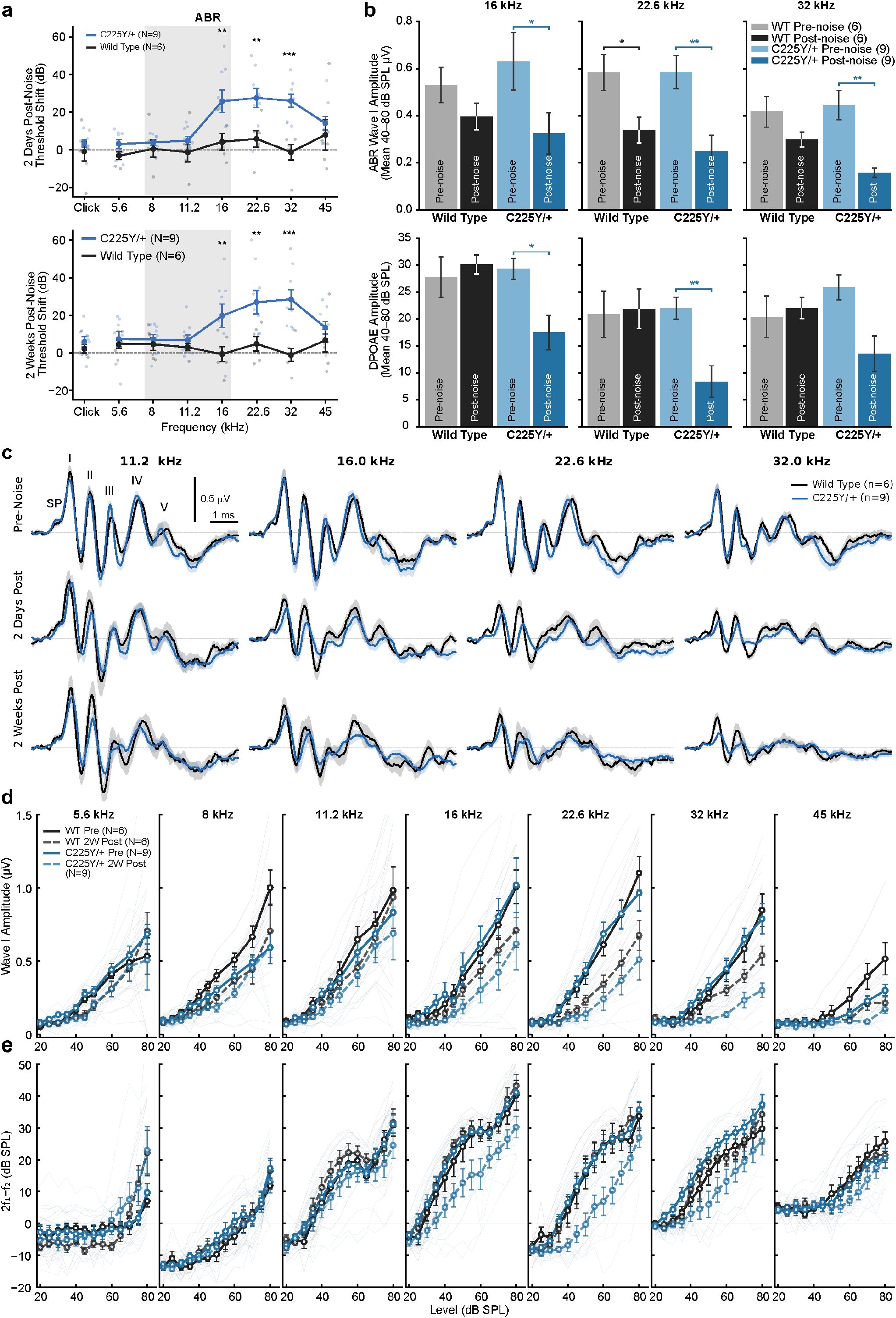
16-week-old *Tectb^C225Y/+^* mice show increased vulnerability to noise damage. (**a**) ABR threshold shifts at 2 days (*top*) and 2 weeks (*bottom*) post-noise exposure. Wild-type and *Tectb^C225Y/+^* mice were exposed for 2 hours to 8–16 kHz octave-band noise at 100 dB SPL (indicated by the shaded area). Threshold shifts were calculated by subtracting pre-exposure values from those obtained on retest after recovery time. When no response was detected at 80 dB SPL, the threshold was entered as 85 dB, one step above the system’s test ceiling. *Tectb^C225Y/+^* mice showed significantly larger threshold shifts with less recovery following noise insult. (**b**) Mean ABR wave I amplitude (top) and DPOAE amplitude (bottom), averaged across the 40–80 dB SPL stimulus range, shown as bar graphs at 16, 22.6, and 32 kHz before and two weeks after noise exposure. (**c**) Average ABR waveforms from wild-type and *Tectb^C225Y/+^* mice recorded before exposure, 2 days after, and 2 weeks after exposure, at 11.2, 16, 22.6, and 32 kHz. *Tectb^C225Y/+^*waveforms show amplitude loss and apparent wave I latency increase post-noise. (**d**) and (**e**), Average growth functions for ABR wave I (*d*) and DPOAE (*e*) before and two weeks after noise exposure. Data are shown as mean ± SEM. *Statistical analysis*: (*a, d, e*) two-way ANOVA followed by Šidák’s multiple comparisons test between genotypes at each frequency; (*b*) Wilcoxon signed-rank paired test. *p < 0.05, **p < 0.01, ***p < 0.001.

**Figure 8.**
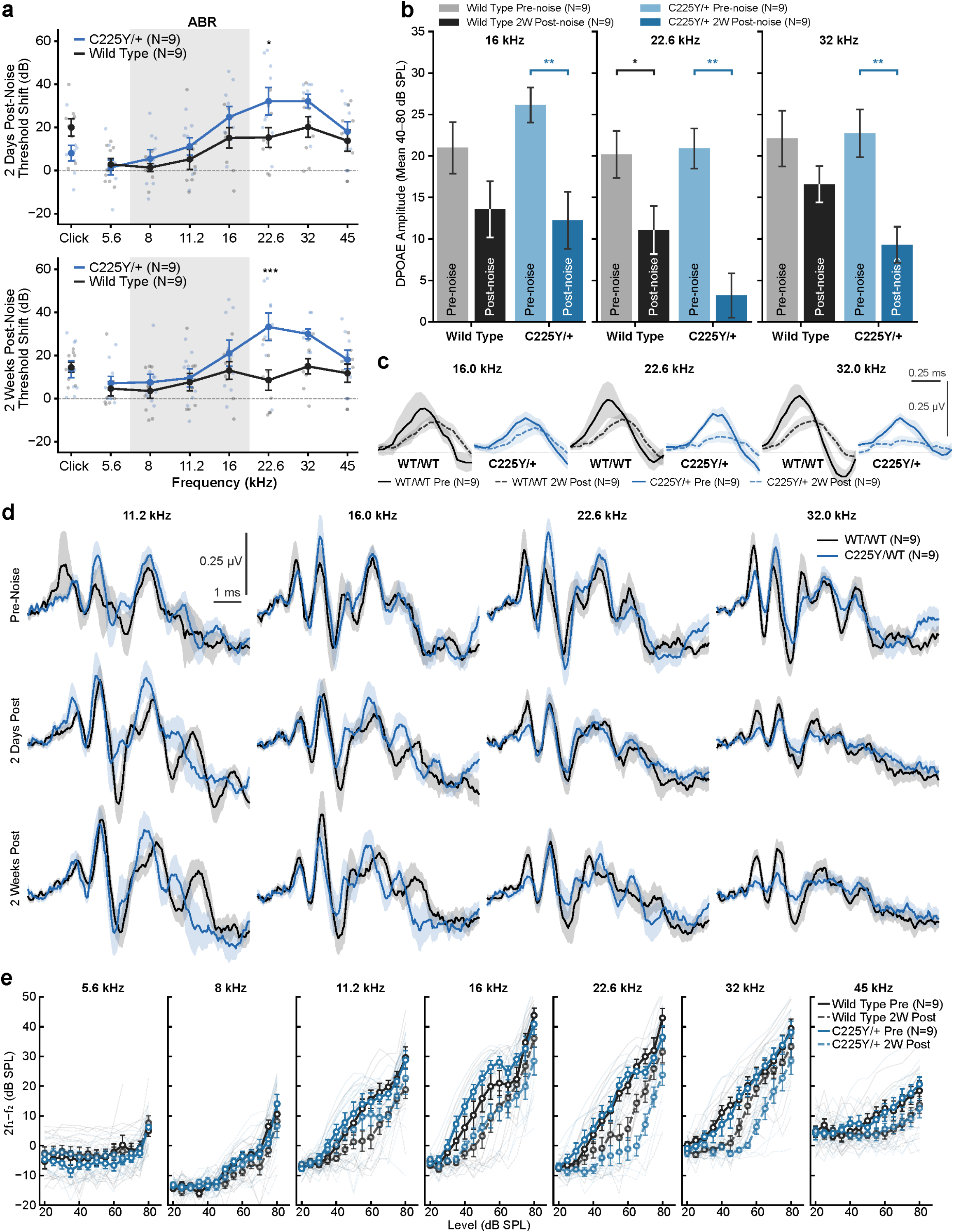
18-month-old *Tectb^C225Y/+^* mice show increased vulnerability to noise damage. (**a**) ABR threshold shifts at 2 days (*top*) and 2 weeks (*bottom*) post-noise exposure. Wild-type and *Tectb^C225Y/+^* mice were exposed for 2 hours to 8–16 kHz octave-band noise at 100 dB SPL (indicated by the shaded area). Threshold shifts were calculated by subtracting pre-exposure values from those obtained on retest after recovery time. When no response was detected at 80 dB SPL, the threshold was entered as 85 dB SPL, one step above the system’s test ceiling. *Tectb^C225Y/+^* mice showed significantly larger threshold shifts with less recovery following noise insult. (**b**) DPOAE amplitude averaged across the 40–80 dB SPL stimulus range, shown as bar graphs at 16, 22.6, and 32 kHz before and two weeks after noise exposure. ABR wave I growth function analysis was not performed in this cohort due to insufficient signal quality of individual waveforms. (**c**) Average ABR waveforms from wild-type and *Tectb^C225Y/+^* mice recorded before and two weeks after noise exposure, shown in the Wave I time window (1.0–1.8 ms) at 16, 22.6, and 32 kHz. Pre-exposure traces (solid lines) and two-week post-exposure traces (dashed lines) are superimposed and presented separately for each genotype. *Tectb^C225Y/+^* mice show greater reduction in Wave I amplitude two weeks following noise exposure compared to wild-type littermates. Data are shown as mean ± SEM, N = 9 per genotype. (**d**) Average ABR waveforms from wild-type and *Tectb^C225Y/+^*mice recorded before exposure, 2 days after, and 2 weeks after exposure, at 11.2, 16, 22.6, and 32 kHz. *Tectb^C225Y/+^* waveforms show amplitude loss and apparent wave I latency increase post-noise. (**e**) Average DPOAE growth functions before and two weeks after noise exposure. Data are shown as mean ± SEM. *Statistical analysis*: (*a, d*) two-way ANOVA followed by Šidák’s multiple comparisons test between genotypes at each frequency; (*b*) Wilcoxon signed-rank paired test. *p < 0.05, **p < 0.01, ***p < 0.001.

To assess OHC function, we next analyzed DPOAE peak amplitude growth as a function of sound intensity across frequencies before noise exposure and at two weeks post-exposure (Figure 7b**,e** and **8b,e**). Panels in Figures 7b and **8b** show average DPOAE amplitudes across the 40–80 dB SPL range for 16, 22.6 and 32kHz as bar graphs, providing a simplified summary of the pre- and two weeks post-noise changes alongside the full growth functions in Figures 7e and **8e**. Prior to exposure, wild-type and *Tectb^C225Y/+^*mice exhibited similar growth functions, indicating comparable OHC-driven distortion product generation under baseline conditions. Following noise exposure, WT mice in the 16-week-old cohort showed recovery of DPOAE amplitudes by two weeks (Figure 7b**,e**), whereas those in the 18-month-old cohort did not (Figure 8b**,e**). *Tectb^C225Y/+^* mice exhibited a persistent depression of the growth function with larger loss over the same interval in both age groups.

To investigate whether this increased susceptibility is accompanied by structural changes in the TM, we analyzed inner ears collected from 18-month-old wild-type and *Tectb^C225Y/+^* mice used in the noise exposure experiments described above (**Figure S9**). We focused this analysis on the older cohort because we reasoned that any genotype-dependent structural consequences of acoustic trauma would be more readily detectable in older animals, where the cumulative effects of aging and noise exposure might be expected to exacerbate subtle TM abnormalities. Using the same light microscopy and quantification pipeline as in Figure 4, we found no significant differences in cross-sectional area or toluidine blue staining density between the two genotypes (**Figure S9b,c**). These results suggest that the functional vulnerability is not accompanied by further structural disorganization detectable at the light microscopy level. Notably, TEM analysis of the 4 kHz cochlear region in 18-month-old mice revealed an absence of SSM in both wild-type and *Tectb^C225Y/+^* animals (**Figure S9d**), consistent with previous reports that SSM is lost or substantially remodeled with normal aging (Bullen et al., 2020; Goodyear et al., 2019).

## Discussion

This study identifies *TECTB* as a human deafness gene and reveals a previously unrecognized role for β-tectorin in providing the cochlea with resilience to age-related and noise-induced hearing loss in mice. We identified two rare heterozygous missense *TECTB* variants segregating with autosomal-dominant, non-progressive SNHL in unrelated multigenerational families. The first, c.674G>A, p.Cys225Tyr *TECTB* variant, identified in multigenerational family 1, affects a highly conserved cysteine residue within the ZP domain, is predicted to be deleterious by multiple *in silico* tools and is absent from population databases. The second, c.853C>T, p.Arg285Cys heterozygous missense variant was identified in an independent family with AD NSHL. This variant is located two amino acid residues from the predicted C-terminal end of the ZP domain and alters a highly conserved residue within the predicted tetrabasic furin-like endoprotease cleavage site between Arg285 and Leu286. Proteolytic cleavage at this site is required for normal folding and polymerization of ZP-domain proteins (Bokhove & Jovine, 2018; Jovine et al., 2005). The variant is predicted to be deleterious and segregates with hearing loss in the six affected individuals. The segregation of two rare, predicted deleterious *TECTB* variants in independent families provides compelling human genetics evidence linking *TECTB* to AD NSHL. Notably, C225Y and R285C are predicted to be mechanistically distinct — disrupting a conserved disulfide bond and a furin cleavage site, respectively — yet both cause autosomal dominant hearing loss, underscoring that ZP domain integrity is broadly required for normal β-tectorin function regardless of the specific molecular perturbation.

To validate the pathogenicity of the c.674G>A, p.Cys225Tyr substitution *in vivo,* we generated a knock-in mouse model carrying the orthologous substitution. The *Tectb-C225Y* allele exhibited a gene-dosage effect. *Tectb^C225Y/C225Y^*mice developed profound deafness with gross ultrastructural deficits of the TM, including complete loss of the SSM. These features resemble the structural phenotype of *Tectb* knockout mice (Russell et al., 2007), indicating that the TECTB-C225Y substitution severely compromises β-tectorin function. In contrast, *Tectb^C225Y/+^*mice maintained normal auditory thresholds under standard housing conditions despite a reduction in TM matrix staining density and subtle morphological abnormalities of key TM features such as the marginal band and Hensen’s stripe. These structural abnormalities, despite the absence of measurable hearing loss, suggest that the variant compromises TM architecture before overt functional deficits become apparent. These findings indicate that under standard laboratory housing conditions, the *Tectb-C225Y* allele alone is insufficient to cause measurable auditory dysfunction in mice, a finding at odds with the human phenotype where hearing loss is evident in heterozygous carriers.

This discrepancy between the human and mouse phenotypes is not without precedent. Several gene discovery studies describing human AD NSHL have identified pathogenic variants that produce an overt phenotype in mouse models only after biallelic disruption (Azaiez et al., 2015; Yan et al., 2013; Zhang et al., 2020). Nevertheless, both intrinsic inter-species differences and environmental factors may contribute to the apparent discrepancy between the human and mouse phenotypes. Several species-specific biological factors may underlie the reduced phenotypic severity in mice, including differences in TM composition, cochlear mechanics, and lifespan. For example, the molecular composition and organization of the TM vary across species (Goodyear & Richardson, 2018), which may influence the functional consequences of structural TM alterations, especially if the relative abundance of specific TM proteins, including TECTB, differs between rodents and humans. In addition, the mechanical coupling between the TM and hair bundles may differ between species. Recent work demonstrated that IHC stereocilia bundles are directly coupled to the TM in guinea pigs (Hakizimana & Fridberger, 2021). However, whether a similar mode of coupling exists in mice or humans remains uncertain. Such species-specific differences in TM–hair bundle interactions could influence how structural TM defects translate into functional auditory deficits. Finally, species-specific compensatory mechanisms or differences in the relative contribution of β-tectorin to TM assembly may further mitigate the functional consequences of the variant in mice compared with humans, although these possibilities remain to be investigated.

To specifically test the environmental component of this discrepancy, we considered the markedly different acoustic environments experienced by humans and laboratory mice. Humans are routinely exposed to a wide range of sound environments throughout life, while laboratory mice are typically housed in relatively quiet, controlled conditions. We therefore hypothesized that noise exposure might unmask latent cochlear vulnerability in *Tectb^C225Y/+^* mice. Indeed, after exposure to a noise trauma paradigm that caused TTS in wild-type mice, *Tectb^C225Y/+^* mice developed significantly larger and permanent ABR threshold shifts, likely recapitulating aspects of the human phenotype. These findings reveal a gene–environment interaction in which the *Tectb-C225Y* variant sensitizes the cochlea to noise insult, leading to long-term functional consequences.

Together, these data suggest that the enhanced noise vulnerability in *Tectb^C225Y/+^* mice reflects impaired cochlear resilience and recovery following acoustic stress, rather than differences in baseline OHC function, and are consistent with compromised TM integrity contributing to increased susceptibility to noise-induced injury. Overall, these findings help reconcile the human–mouse phenotype discrepancy and suggest that the *TECTB-C225Y* variant creates a structurally vulnerable TM that functions adequately under baseline conditions but renders the cochlea more susceptible to environmental noise. While the acute noise paradigm used in our study effectively revealed this latent susceptibility to acoustic injury, future studies employing chronic or intermittent low-intensity noise exposure paradigms (Feng et al., 2020; Maison et al., 2013) will help determine whether more physiologically relevant acoustic environments similarly unmask progressive hearing deficits in *Tectb^C225Y/+^* mice.

Post-noise ABR wave I growth functions were reduced in *Tectb^C225Y/+^*mice relative to wild-type at both ages examined, and DPOAE growth functions were similarly depressed. This contrasts with the selective ABR deficit observed in intact *Tectb^C225Y/C225Y^* mice and suggests that a TTS-level noise exposure in heterozygous animals with already-compromised TM architecture produces broad cochlear dysfunction through impaired stimulus transmission to, or amplified mechanical stress on, OHC and IHC stereocilia bundles that is sufficient to result in a PTS.

The disruption of the ZP domain in TECTA is characterized as mid- or pan-frequency hearing impairment in patients with DFNA8/12 (Verhoeven et al., 1998). These individuals present highly variable audiogram profiles including mild-to-severe and have an age of onset ranging from early childhood to adolescence (Chen et al., 2022; Yasukawa et al., 2019). Multiple studies have reported both progression (Moteki et al., 2012) and non-progression (Iwasaki et al., 2002) of hearing loss for patients with variants disrupting the ZP domain. In those with progressive hearing loss, deterioration was not accelerated by particular TECTA variants but rather by age (Yasukawa et al., 2019).

The *TECTB-C225Y* substitution occurs at a highly conserved cysteine within the ZP domain, a motif critical for the polymerization and stabilization of many extracellular matrix proteins (Bokhove & Jovine, 2018). The absence of the SSM in *Tectb^C225Y/C225Y^* mice, along with our indirect measures reporting reduced TM matrix density in both *Tectb^C225Y/+^*and *Tectb^C225Y/C225Y^* mice, suggests that the variant interferes with the correct incorporation of β-tectorin into the TM matrix and/or its stability once incorporated, leading to functional deficit. These observations align with prior studies of TECTA mutations (Chen et al., 2022; Yasukawa et al., 2019), some of which are also predicted to affect ZP-domain interactions and lead to dominant or recessive hearing loss depending on the nature and position of the variant (Iwasaki et al., 2002; Moteki et al., 2012). Notably, pathogenic TECTA ZP-domain variants often produce mid-frequency or flat hearing loss with variable progression, similar to the audiometric phenotype observed in our patient cohort, and have also been associated with TM structural abnormalities resembling those we observed in *Tectb^C225Y/+^* mice (Legan et al., 2014; Legan et al., 2005; Xia et al., 2010).

Audiometric data from family 1 showed mostly stable hearing loss ranging from mild to severe, even among siblings, and no evidence of progression over time, as confirmed by re-testing three family members over a decade after the initial hearing test was administered, strengthening confidence in the non-progressive nature of the phenotype. Family 2 showed two audiogram configurations; namely, sloping high frequency hearing loss as seen in individuals II:5, III:4, and III:4 at 56-60, and 36-40 years of age at last measurement, respectively. The younger individual (III:5) so far shows mild hearing loss with a distinct 4 kHz notch that is maintained in over 10 years of audiometry. This variability in phenotype expression despite a shared genotype supports a model in which environmental modifiers, particularly noise exposure, are likely to contribute to disease manifestation. Our findings in *Tectb^C225Y/+^* mice provide direct experimental support for this hypothesis and suggest that the TECTB-C225Y variant creates a sensitized cochlear environment—likely to be functionally silent under ideal conditions but prone to early decompensation under stress.

In addition to the direct reporting on the *TECTB-C225Y* variant, our results highlight the importance of mouse strain genetic background in auditory phenotyping. Earlier studies of TM protein mutants, including *Tecta* knock-ins, were constrained by seizure susceptibility in certain mouse strains, which precluded noise exposure experiments (Legan et al., 2014). By generating the *Tectb-C225Y* mouse model on a C57BL/6J background with a corrected *Cdh23^Ahl+^* allele, we avoided these limitations and enabled the interrogation of both baseline and environmentally induced phenotypes following noise insult. The ability to model gene-environment interactions in this setting was critical for uncovering the additional effects conferred by the *TECTB-C225Y* variant and may be relevant to other deafness genes whose phenotypic impact might also be conditional on environmental stressors that remain untested.

The distinction between ABR and DPOAE responses in young, unexposed *Tectb^C225Y/C225Y^*mice is an unexpected finding that provides mechanistic insight. While DPOAEs reflect the electromotility of OHCs, ABRs integrate responses across the full auditory pathway, from cochlear mechanics to neural signaling. Thus, largely preserved DPOAEs alongside more severely impaired ABRs suggest that the functional deficit in *Tectb^C225Y/C225Y^* mice is more pronounced at the level of IHC stimulation than OHC function. This interpretation is further supported by the unchanged relative contribution of the summating potential and the absence of overt synaptic abnormalities which argue, respectively, against an impairment of mechano-transduction and/or synaptopathy as principal explanations for the relatively larger ABR defects. Instead, these findings support a model in which disruption of the TM compromises the mechanical drive to IHC stereocilia while largely preserving OHC bundle stimulation. Given that TECTB localization is restricted to the TM, this interpretation provides the most parsimonious explanation for the disproportionately larger reduction in ABR responses observed in *Tectb^C225Y/C225Y^*mice. Consistent with this interpretation, the TM was the only cochlear structure in which we detected consistent morphological abnormalities, with structural disorganization and reduced matrix staining density in *Tectb^C225Y/C225Y^*mice, consistent with impaired TM integrity as the primary anatomical substrate underlying the functional deficit.

Dissociation between ABR and DPOAE responses has been reported previously in experimental conditions involving altered cochlear mechanics (Puria et al., 1996). Computational modeling predicts that changes of cochlear fluid viscosity have a greater impact on fluid-coupled IHC stereocilia while having a smaller impact on OHC bundles that remain directly coupled to the TM (Wang et al., 2016). This is consistent with previous observations in *Tecta*-*Y1870C* and *Otoa-EGFP* knock-in mice, which demonstrated a critical role for the TM in driving IHC stimulation, with isolated TM defects leaving OHC function largely unaffected (Legan et al., 2005; Lukashkin et al., 2012). Conversely, when the TM becomes detached from OHC bundles, OHCs themselves become fluid-coupled, as evidenced by a phase shift in cochlear microphonic responses observed in *Tecta^ΔENT/ΔENT^* and *Tecta/Tectb* double-knockout mice, demonstrating that TM structural integrity governs the mode of stimulation of both hair cell types (Jeng et al., 2021; Legan et al., 2000). Particularly relevant to the present study, *Tecta^C1509G/+^* mice, which carry a shortened TM that contacts only the first row of OHCs while exhibiting normal DPOAE thresholds under quiet conditions, show incomplete threshold recovery and increased OHC loss following noise exposure. Computational modeling predicted approximately 50% higher shear forces on OHC stereocilia in these mice, suggesting that structural abnormalities of the TM can amplify mechanical stress on hair-cell stereocilia during acoustic exposure and increase susceptibility to noise-induced injury (Liu et al., 2011).

While TECTB immunolabeling in the cochlea has only been observed in the TM (Russell et al., 2007), suggesting that its role in hearing is primarily localized to this structure, we cannot entirely exclude the possibility of additional or indirect functions elsewhere in the cochlea. However, the greater ABR and DPOAE deficits observed in noise-exposed *Tectb^C225Y/+^* mice compared to wild-type littermates are consistent with impaired cochlear resilience and support the interpretation that a weakened TM architecture renders the cochlea more susceptible to noise insult.

Our data support a semidominant model for the mouse *Tectb-C225Y* allele. *Tectb^C225Y/C225Y^* mice display a severe, early-onset hearing loss with gross TM deficits. *Tectb^C225Y/+^* mice exhibit structural TM abnormalities and a functionally silent phenotype under baseline conditions but display an increased sensitivity to noise insult. The reduced matrix staining density and structural abnormalities of the TM caused by the TECTB-C225Y variant are likely to alter the viscoelastic properties of the TM or its interactions with hair-cell stereocilia. This model echoes prior findings from *Tecta* ZP-domain mutants, where dominant phenotypes often arise from structural disorganization rather than protein loss *per se*. Furthermore, it supports the notion that subtle TM disruptions in humans may contribute to complex hearing loss phenotypes that are currently underdiagnosed, particularly those with unexplained increased noise sensitivity or age-related declines. Future studies combining direct measurements of TM biomechanics with assessment of IHC function will be important for defining how altered TM mechanics impair cochlear resilience and produce the characteristic dissociation between ABR and DPOAE responses observed in *Tectb^C225Y/C225Y^* mice.

Notably, TEM analysis revealed that the SSM is absent in both wild-type and *Tectb^C225Y/+^* animals at 18 months of age, consistent with normal age-related SSM remodeling (Bullen et al., 2020; Goodyear et al., 2019). The persistent noise vulnerability in aged *Tectb^C225Y/+^* mice therefore cannot be attributed to SSM loss alone and likely reflects more broadly compromised TM architecture — including marginal band abnormalities and reduced matrix density — that precedes and outlasts the age-related loss of SSM seen in both genotypes.

Although we did not directly assess the TM’s biophysical properties, our electrophysiological and morphological findings are consistent with altered extracellular matrix architecture and impaired function. The specific interactions of β-tectorin with other TM proteins remain to be defined, but future studies leveraging structural biology and protein chemistry may elucidate the relevant binding interfaces and clarify how disease-associated variants disrupt matrix assembly.

Taken together, this study provides evidence that *TECTB* is a *bona fide* human deafness gene and demonstrates that a missense variant within its ZP domain compromises TM integrity and cochlear resilience to noise insult. The findings also illustrate the value of cross-species modeling and the importance of considering environmental context in interpreting genotype–phenotype relationships. Finally, these results argue for the inclusion of *TECTB* in diagnostic panels for hereditary hearing loss.

## Materials and Methods

### Family recruitment and clinical assessment

This study was approved by the institutional review boards of Dartmouth-Hitchcock Medical Center (No. 22289) and the Nicaraguan Ministry of Health and by the ethics committee at the Medical Faculty of the University of Würzburg (approval numbers 131/21, 226/18 and 46/15).The proband in family 1 was enrolled in a study focusing on the genetic etiology of hereditary hearing loss rural communities. The proband in family 2 was enrolled in an undiagnosed hearing loss research study. Written informed consent was obtained from participating members from families 1 (II:1, II:2, II:3, II:4, II:5, III:1, III:2, III:3, III:4, III:5, III:6, and IV:1) and 2 (I:2, II:2, II:4, II:5, II:6, III:4, and III:5). Genomic DNA of participating family members was extracted from peripheral blood leukocytes.

Participating individuals from family 1 completed a risk factor questionnaire that reviewed perinatal and birth history, neonatal or perinatal infections, maternal exposure to rubella, exposure to ototoxic drugs or pesticides, other congenital or significant medical problems, ear infection history, and noise exposure. Detailed physical examination included an otoscopic and craniofacial examination and assessment of dysmorphic features including the craniofacial development and examination of the pinna, otoscopic examination of the ear, and examination of other physical features known to be associated with syndromic hearing loss (ocular exam, skin pigmentation abnormalities, limb abnormalities). Audiometric data included pure-tone thresholds for 0.5, 1, 2, 4, and 8 kHz, which were collected as previously reported (Saunders et al., 2007). Both air and bone thresholds were obtained. Severity of hearing loss was determined by averaging pure-tone thresholds over 0.5, 1, 2, and 4 kHz (PTA0.5-4kHz) for the better ear. The severity of hearing loss in the better ear was defined as mild for thresholds averaging 20-40 dB hearing level (HL), moderate for 41-70 dB HL, severe for 71-95 dB HL, and profound in excess of 95 dB HL. Progressive hearing loss was defined as a deterioration of >15 dB HL in the average over the frequencies of 0.5, 1, and 2 kHz within a 10-year period. Otoacoustic emissions were assessed in individuals II:4, II:5, and III:2 from family 1 and individual III:5 in family 2.

### Genetic testing of family 1

High-throughput sequencing using the OtoGenome® 74-gene panel (**Table S4**) was performed on the proband’s DNA sample using oligonucleotide-based target capture (Agilent SureSelect) followed by Illumina HiSeq sequencing. Variant calls were generated using the Burrows-Wheeler Aligner followed by GATK analysis. Variants were mapped to human genome reference build hg19. This test detected >99% of substitution variants (95% CI = 98.5-100) and 95% of small insertions and deletions (95% CI = 83.1 - 98.6). Following an uninformative result, the proband’s sample was subjected to 2 x 75 bp paired-end exome sequencing v3 (Broad Institute, Cambridge, MA, USA). Variant calls were annotated by Annovar (Wang et al., 2010).

### Genotyping, copy number variation calling and linkage analysis

Whole genome genotyping using the genomic DNA samples from family 1 was performed using the HumanOmni2.5-8v1 SNP array (Illumina, San Diego, CA, USA) at Partners Research Core. Copy number variation (CNV) calls were visualized using GenomeStudio following calling with pennCNV and CNVPartition (Illumina). Mendelian errors due to genotype errors were checked with Pedcheck and erroneous genotypes were removed (O’Connell & Weeks, 1998). A parametric model was used, as the pedigrees showed AD inheritance. Multipoint linkage analysis was performed using Allegro version 2.0 (Gudbjartsson et al., 2005). Fine mapping was performed of the chromosome 10 linkage peak.

### Genetic testing of family 2

Two separate sequencing approaches were conducted. The genomic DNA of the proband (II:5), unaffected partner (II:6) and affected child (III:5) were sequenced with the TruSight One panel (Illumina) containing 4,813 genes. Exome sequencing was later performed on the genomic DNA of the proband (II:5), his affected sibling (II:2) and affected child (III:5) using the Exome Nextera Rapid Kit (Illumina) and 2 x 75 bp paired-end sequencing. Data analysis was done using GenSearch NGS as previously described (Hofrichter et al., 2018).

### Variant assessment and validation

Exome data were filtered to remove variants with an allele frequency of 0.01 or more in the population frequency databases gnomAD v4.1.0 and *All of Us Data Browser* (All of Us Research Program Genomics, 2024; Chen et al., 2024) and variants with an allelic balance less than 20%. Coding and splice site variants were retained for analysis. Deleteriousness of missense variants was assigned according to the prediction from multiple prediction tools (SIFT, PolyPhen-2, FATHMM, MutationTaster, REVEL, CADD, and ClinPred) and supported by evolutionary conservation of the affected amino acid as assessed by phyloP. Splicing predictions utilized the splice tools embedded in Alamut Visual Plus as well as Splice AI Visual (de Sainte Agathe et al., 2023). Both heterozygous and homozygous variants were analyzed. Variants suspected of having potential clinical significance related to the probands’ hearing loss were confirmed by Sanger sequencing in family members to assess segregation with the disorder. Sanger sequencing was performed in family 1 (*TECTB* Exon 8 F: 5’-CTCGGGAGCCTTGTGTGTAA-3’ and *TECTB* Exon 8 R: 5’-CTCCTCCAATACTGGCACCC-3’) and family 2 (*TECTB* Exon 9 F: 5’- GGATGAACAGTGTGTCTTCTGG-3’ and *TECTB* Exon 9 R: 5’- GTTTGGCTTGAGCTGTTCC-3’) using standard protocols.

### Generation of the *Tectb-C225Y* knock-in mouse line

All procedures and protocols were approved by the Institutional Animal Care and Use Committees of Mass Eye and Ear and of the University of Nebraska Medical Center. For work conducted at the University of Sussex, animals were bred and maintained in accordance with UK Home Office guidelines, and the experiments were performed in accordance with the Home Office Animals (Scientific Procedures) Act 1986, following approval by the Animal Welfare Ethical Review Board at the University of Sussex. All aspects of animal care, use, and experimental procedures were conducted in compliance with relevant institutional and national ethical regulations governing animal research.

#### CRISPR strategy

The *Tectb-C225Y* mouse line was created using the CRISPR-Cas9 system. The guide RNA sequence used for changing the codon TGC to TAC was TGTCTCATCGGTGGG **GC/A***GCTGG*. The guide is in antisense orientation. Codon 225 (TGC) is bold-faced, and the PAM sequence is shown in italics. The guide cut site is shown with the symbol “/”.

#### CRISPR reagents

The guide RNA was synthesized by annealing two ultramer oligonucleotides (atatcggatcccTAATACGACTCACTATAGGTGTCTCATCGGTGGGGCAGcGTTTTAGAGCTAGAAATAGCAAGTTAAAATAAGGCTAGTCCGTTATCAACTTGAAAAAGTGGCACCGAGTCGGTGCTTTTTT and AAAAAAAGCACCGACTCGGTGCCACTTTTTCAAGTTGATAACGGACTAGCCTTATT TTAACTT GCTATTTCTAGCTCTAAAACgCTGCCCCACCGATGAGACACCTATAGTGAGTCGTATTAgggatccga tat) followed by in vitro transcription using MEGAshortscript T7 kit (Ambion: AM 1354). The Tectb repair DNA (ultramer) sequence was GAGCCTTTTTGGAGCTCTCTGGTGTGATGTTT CCTTCTCCCTTCC CTCTGCATCCCAGC<u>T**A**C</u>CCCACCGATGAGACAGTCCTCGTGCATGAGAACGGCAAAGACCACA GGGCCACTTTCCA. The ultramers were purchased from IDT (Coralville, Iowa, USA). Codon 225 Y (TAC) is underlined and the nucleotide incorporated is shown in bold-faced letter. The Cas9 mRNA was synthesized using the pBGK plasmid (Harms et al., 2014; Miura et al., 2018) as a template for invitro transcription using mMESSAGE mMACHINE T7 ULTRA kit (Ambion: AM 1345).

#### Pronuclear injection

C57BL/6 mice were used as embryo donors. Detailed descriptions of CRISPR/Cas9-mediated mouse genome editing are described in (Quadros et al., 2015). Briefly, the injection mix contained 10 ng/μL each of sgRNA, repair-DNA ultramer (ssDNA) and Cas9 mRNA.

#### Genotyping of offspring and nucleotide sequencing

Genomic DNAs were extracted from the offspring using Qiagen Gentra Puregene Tissue Kit and the target region was PCR-amplified using primers F1 (CATCCATTCCCGTGGCTCTG) and R1 (GACTGGAGGAGCCGGATTTG). The wild-type sequence contains a *PvuII* Restriction Endonuclease site and successful homologous recombination results in the loss of a *PvuII* site. The PCR products were genotyped by Restriction Fragment Length Polymorphism (RFLP, see **Figure S3a**). Primers F2 (GCTCCTCACAAGCCTAGCAG) and R2 (GCGACAGTGAGAAGCTCTCC) were then used to amplify products that were Sanger sequenced with the R2 primer to confirm presence of the mutant allele (**Figure S3b**).

The resulting *Tectb-C225Y* allele (JAX Stock No. 041413; B6(CAST)-Cdh23<Ahl> Tectb<Em1Guru>/GprIndzJ, MGI:8277171) was then transferred to a C57BL/6J-*Cdh23^Ahl+^* strain (Jax stock # 002756) which retains the line’s background while correcting the *Cdh23^Ahl^* mutation known to cause age-related hearing loss in the C57BL/6J line. Test groups consisted of mixed male and female mice from shared large litters, which were aged for testing and tissue collection at pre-determined time points on site. This approach ensures standardized experimental conditions and evaluates both genders to control for any potential sex-specific effects. Animals were randomly allocated to experimental groups.

### Auditory testing

ABR and DPOAE measurements were performed in a sound-attenuating and electrically shielded chamber using the *EPL Cochlear Function Test Suite* (EPL Acoustic System, Eaton-Peabody Laboratories) at Mass Eye and Ear. Recordings were collected at six time points: 5 weeks, and 3, 6, 9, 12, and 18 months of age. Each mouse underwent both ABR and DPOAE testing in a single session per time point.

Mice were anesthetized with an intraperitoneal injection of ketamine (120 mg/kg) and xylazine (10 mg/kg). Artificial tears were applied to prevent corneal drying. Body temperature was maintained at 37 °C throughout the procedure using a thermostatically controlled heating pad. Tympanic membranes were visually inspected for obstructions or anomalies prior to testing. Subdermal needle electrodes were inserted at the vertex (non-inverting), post-auricularly behind the stimulated ear (inverting), and near the base of the tail (ground).

ABRs were recorded in response to both broadband clicks and pure tone stimuli at 5.6, 8, 11.2, 16, 22.6, 32, and 45 kHz. Each tone pip was 5 ms in duration, with a 0.5 ms rise-fall time, delivered in alternating polarity at a rate of 30 stimuli per second. Stimulus intensity ranged from 20 to 80 dB SPL in 5 dB steps. Responses were amplified 10,000×, filtered between 0.3 and 3 kHz, and averaged across 512 repetitions, as previously reported (Strelkova et al., 2024). ABR thresholds were determined using the *ABR Peak Analysis* tool (Suthakar & Liberman, 2019), then visually inspected and corrected as needed. ABR thresholds were defined as the lowest stimulus intensity that evoked repeatable and identifiable waveform peaks.

DPOAEs were recorded in response to two simultaneous pure-tone stimuli, *f1* and *f2* (f2/f1 = 1.2), where *f2* frequencies matched those used in ABR testing (5.6, 8, 11.2, 16, 22.6, 32, and 45 kHz). Primary tone levels were swept from 10 to 80 dB SPL for f2 in 5 dB increments, with L1 set to L2 + 10 dB. DPOAEs were recorded at the 2*f1*–*f2* frequency using a low-noise microphone mounted within the APL acoustic system assembly. Iso-response contours were interpolated from plots of 2*f1*–*f2* amplitude versus *f1* level. The DPOAE threshold was defined as the f1 level required to elicit a 2*f1*–*f2* distortion product of 0 dB SPL. DPOAE thresholds were reported only for 8, 11.2, 16, 22.6, and 32 kHz, as measurements at 4, 5.6, and 45 kHz were highly variable across groups, likely due to equipment limitations and calibration inconsistencies at these edge frequencies.

In cases when the strongest sound stimulus (80 dB SPL) resulted in no detectable response, the threshold was reported as 85 dB SPL and indicated with an upward-pointing arrow on the graphs. All responses were then analyzed and plotted in *GraphPad Prism* or using *Python* plotting functions. In addition to thresholds, ABR peak I amplitude growth functions and latencies were analyzed for certain age groups and genotypes as reported in the manuscript.

Analysis scripts for ABR waveform extraction, data organization, and statistical analyses contributing to Figures 2, 3, 7, 8 and S4 were developed in *Python* with assistance from *Claude Sonnet 4.6* (*Anthropic*; accessed 2026). All AI-assisted code was reviewed, tested, and revised by the authors, who verified accuracy and reproducibility of all results. The authors independently designed all analytical workflows, including data inclusion criteria, grouping variables, and statistical methods. To further validate the findings, all key results were independently reproduced using manual data pooling and analysis in *GraphPad Prism*. The authors take full responsibility for all analyses and conclusions presented in this study.

### Noise exposure

Test cohorts were established at two time points: a young age of 16 weeks and an advanced age of 18 months, utilizing mice that had recently undergone longitudinal auditory testing. Following baseline ABR and DPOAE assessments (as described above), awake, unrestrained mice were placed individually in small cages and exposed to octave-band noise (8–16 kHz) for 2 hours at 100 dB SPL to induce TTS in the age groups used in our study (Jensen et al., 2015; Kujawa & Liberman, 2006). The noise was generated from a white-noise source, filtered, amplified, and delivered via a horn mounted atop a reverberant tabletop exposure booth. Exposure levels were confirmed in each cage using a 0.25-inch Brüel and Kjær condenser microphone.

ABRs and DPOAE thresholds were then reassessed 2 days and 2 weeks after exposure to evaluate auditory recovery. Threshold shifts from pre-exposure levels were calculated for each frequency and time point in each animal. Three animals were excluded from the analysis (one WT and one *Tectb^C225Y/+^* mouse at 16 weeks, and one 18-month-old *Tectb^C225Y/+^* mouse) due to undetectable pre-exposure ABR or DPOAE responses at one or more reported frequencies. DPOAE threshold shifts were reported only for 8, 11.2, 16, and 22.6 kHz, as measurements at 4, 5.6, 32, and 45 kHz were highly variable across groups, likely due to equipment limitations and calibration inconsistencies at these edge frequencies.

### Tissue processing and imaging

Cochleae were removed and placed in petri dishes containing PBS. The oval and round windows were opened, and a small hole was made through the bone at the apical end of each cochlea. A small volume (∼20 μL) of fixative (2.5% glutaraldehyde in 0.1 M sodium cacodylate, pH 7.4 (EMS 15960), containing 10 mg/mL tannic acid (Sigma 403040)) was slowly perfused through the oval window and a further 20 μL of fixative was delivered through the hole at the apical end. Further holes were subsequently made in the lateral walls of the middle and basal turns. Cochleae were then immersed for ∼12 h in fixative at 4°C on a rocker. Following three washes in 0.1 M sodium cacodylate, cochleae were post-fixed in 1% osmium tetroxide (EMS 19150) in 0.1 M sodium cacodylate pH 7.4 for 4 h at room temperature. Cochleae were then washed a further three times in 0.1 M sodium cacodylate and decalcified in 0.5 M EDTA (Millipore 324504) containing 0.1% glutaraldehyde (EMS 16019) for 5 days at 4°C on a rocker. Following a wash in distilled water, cochleae were dehydrated through an ascending ethanol series, equilibrated in propylene oxide and infiltrated and embedded in Epon 812/Araldite 502 resin (EMS). After curing at 60°C for 48-72 h with the cochleae positioned so that the oval and round windows faced upwards, cochleae were mounted with the windows facing downwards and sectioned until profiles of the ∼4, 8, 20, and 40 kHz regions could be obtained from a single section.

For light microscopy, 1 μm sections were stained with 1% (w/v) Toluidine blue containing 1% (w/v) borax, prior to widefield imaging. For TEM, ultrathin sections (∼80 nm) were collected onto formvar/carbon-coated slot grids. Sections were stained on grids with 1% uranyl acetate and lead citrate, then imaged on a Hitachi TH7800 TEM operated at 100 kV at the Center for Nanoscale Systems, Harvard University. Images were processed in Adobe Creative Suite. Ultrathin sections on copper grids were also stained as described and imaged with a JEOL 1400+ TEM operating at 80 kV in the Electron Microscopy Imaging Centre at the University of Sussex. Images were captured with a Gatan Ultrascan 1000+ CCD camera and processed with Adobe Photoshop.

For immunostaining of cryosections, the inner ears were immersion-fixed after removal of the oval and round windows in 3.7% formaldehyde (10% formalin) in 0.1 M sodium phosphate buffer pH 7.4 for 2-3 hours at room temperature, washed 3 x in PBS and decalcified at 4°C in 0.5 M EDTA pH 8.0 for 3 days. After washing 3x with PBS to remove the EDTA, samples were equilibrated with 30% sucrose in PBS, embedded in 1% low-gelling temperature agarose in PBS containing 18% sucrose, mounted onto cryotome chucks coated with a thin layer of OCT compound, fast frozen using Cryospray and cryosectioned at a temperature of ∼-30°C and a thickness of 15 microns. Cryosections were mounted on gelatin-coated glass slides, dried overnight at 37°C, preblocked with 10% heat-inactivated horse serum in PBS (PBS/HS) for 1 hour, and stained overnight at room temperature with rabbit anti-chick TECTB (R7, (Knipper et al., 2001)), rabbit anti-chick TECTA (R9 (Knipper et al., 2001)), rabbit anti-mouse CEACAM16 (Zheng et al., 2011) and rabbit anti-pig COL9A (Richardson et al., 1987) at a dilution of 1:1000. Sections were washed, labelled for 1 hour with Texas-Red conjugated phalloidin (Invitrogen) and Alexa-488 goat anti-rabbit Ig, both at a dilution of 1:500 in PBS/HS, washed in PBS, mounted in Vectashield and imaged using a Zeiss Axioplan2 equipped with a Spot RT camera and a 20ξ lens.

For whole-mount immunostaining and quantification of IHC ribbon synapses, the inner ears from 10-week-old wild-type, *Tectb^C225Y/+^*and *Tectb^C225Y/C225Y^* littermates were fixed and decalcified as described above. After washing 3x in PBS, cochleae were microdissected into half-turns and the middle turn was retained for analysis. Whole mounts were permeabilized and blocked in a single step for 1 hour in PBS containing 0.1% Triton X-100 and 10% goat serum, and stained overnight at 4°C with mouse anti-CtBP2 (BD Transduction Laboratories, Cat #612044, RRID:AB_399431) and rabbit anti-Homer1 (GeneTex, Cat #GTX103278) to label the presynaptic ribbons and postsynaptic densities, respectively, and guinea pig anti-Vglut3 (Synaptic Systems, Cat #135204, RRID:AB_2619825) to label the inner hair cells, all at a dilution of 1:500. Whole mounts were washed and labelled for 2 hours at room temperature with Alexa Fluor Plus 488 goat anti-mouse IgG (Invitrogen, Cat #A32723, RRID:AB_2633275), Alexa Fluor 568 goat anti-rabbit IgG (Invitrogen, Cat #A-11011, RRID:AB_143157) and DyLight 405 goat anti-guinea pig IgG (Jackson ImmunoResearch, Cat #106-475-003, RRID:AB_2337432), all at a dilution of 1:1000 in PBS-goat serum, washed in PBS and mounted in ProLong Antifade mountant.

Confocal Z-stacks were acquired with a Leica SP8 confocal using a 63×, 1.4 NA oil-immersion lens. Juxtaposed CtBP2/Homer1 puncta lying within the inner hair cell volume were counted as synapses.

### Staining and quantification of TM cross-section area

Adaptive thresholding was utilized in order to quantify the cross-sectional area of the TM and density of stained material as described previously (Cheatham et al., 2014). Briefly, while blinded to genotype, the outer border of TMs were traced in Adobe Photoshop CC, and area calculated. A threshold level that excluded all but pixels devoid of stain was identified for each image utilizing the Image J adaptive threshold tool, and the area of these pixels was calculated as a percentage of TMs cross-sectional area.

### Quantification of ultrastructure deficits

Marginal band and Hensen’s stripe morphology in cross-sections of the 40 kHz region was scored in a random order, blind to genotype. Morphology scores were pooled from 3 independent investigators. Marginal bands were scored as 1-normal, 2-mishapen, 3-severely misshapen. Hensen’s stripes were scored as 1-normal, 2-disrupted, 3-absent.

### Statistical Analysis

*TM cross-sectional area and TM unstained area.* One-way ANOVAs were carried out at each frequency with Šidák’s multi-comparison tests between each genotype, followed by Benjamini-Hochberg correction for all comparisons within each age group. Proportions of TM sections devoid of stain (%) underwent arcsin square root transformation prior to statistical analysis in order to better satisfy the requirements of a one-way ANOVA. All statistical analyses were carried out in *GraphPad Prism*, with the exception of Benjamini-Hochberg corrections which were calculated manually.

*ABR and DPOAE Results.* Two-way ANOVAs were carried out in *GraphPad Prism* or *Python* to compare each frequency between genotypes at each age point. Aging data comparing three genotypes and noise exposure data comparing two genotypes were analyzed with Šidák’s multiple-comparisons test, as specified in each figure legend. All statistical analyses were carried out in *GraphPad Prism* or *Python*.

## Acknowledgements

The authors acknowledge Dr. David P. Corey, and Dr. Sharon Vaz for their valuable contributions to this study, as well as the patients and family members who participated in and supported this research. This work was done with the support of the Center for Rare Hearing Disorders at the Center of Rare Diseases Göttingen (ZSEG).

The authors acknowledge the use of *Claude Sonnet 4.6* (*Anthropic*; accessed 2026) to assist with the development of *Python* scripts for data analysis and generation of selected figure panels, and *ChatGPT* (*OpenAI* GPT-5.5; accessed 2026) to assist with revising selected passages of the manuscript. All AI-assisted code and text were reviewed, tested and revised, as appropriate, by the authors. The authors verified the accuracy and reproducibility of all analyses and ensured the accuracy and consistency of the manuscript with the data presented.

## Author contributions

**E.B.H.:** Conceptualization, methodology, validation, formal analysis, investigation, data curation, writing-original draft, writing-review and editing, visualization, funding acquisition; **B.V.:** Validation, formal analysis, investigation, data curation, visualization, writing – original draft, project administration, funding acquisition; **R.J.G.:** Conceptualization, methodology, validation, formal analysis, investigation, data curation, visualization, writing – review & editing; **R.T.O.:** Investigation, methodology, validation, formal analysis, data curation, writing-original draft; **J.Sh.:** Conceptualization, methodology, validation, formal analysis, investigation, supervision, funding acquisition; **S.S.A.:** Resources; **K.M.:** Validation, investigation, data curation; **M.A.H.H.:** Investigation, methodology, validation, formal analysis, data curation; **J.Sc.:** Investigation, data curation; **S.F.:** Investigation; **P.G.:** Investigation, methodology, validation, formal analysis, visualization; **R.V.M.:** Investigation; **K.C.:** Investigation; **K.L.G.:** Investigation; **R.Q.:** Investigation, validation; **M.O.:** Investigation, validation; **J.M.:** Investigation; **E.J.W.:** Investigation; **C.C.M.:** Resources, supervision, project administration; **T.H.:** Resources, supervision; **C.G.:** Resources, validation, formal analysis, supervision; **J.E.S.:** Conceptualization, methodology, investigation, data curation, project administration, supervision, funding acquisition; **G.P.R.:** Conceptualization, methodology, validation, formal analysis, investigation, data curation, writing – original draft, project administration, supervision, funding acquisition; **A.A.I.:** Conceptualization, formal analysis, investigation, data curation, visualization, writing – original draft, project administration, supervision, funding acquisition.

## Data availability

The source data used to generate the graphs in this manuscript are available in the supplementary data file. Further imaging data that support the findings of this study are available from the corresponding authors upon request.

## Ethics approvals

Human studies were approved by the institutional review board of Dartmouth-Hitchcock Medical Center (No. 22289) and the Nicaraguan Ministry of Health and by the ethics committee at the Medical Faculty of the University of Würzburg (approval numbers 131/21, 226/18 and 46/15).

## Consent to participate and publish

Written informed consent was obtained from all participants including permission to publish.

