## Supplementalry Figures for "*TECTB* Variants Reveal Tectorial Membrane Vulnerability in Dominant Non-Syndromic Hearing Loss"

### Supplementary Figures

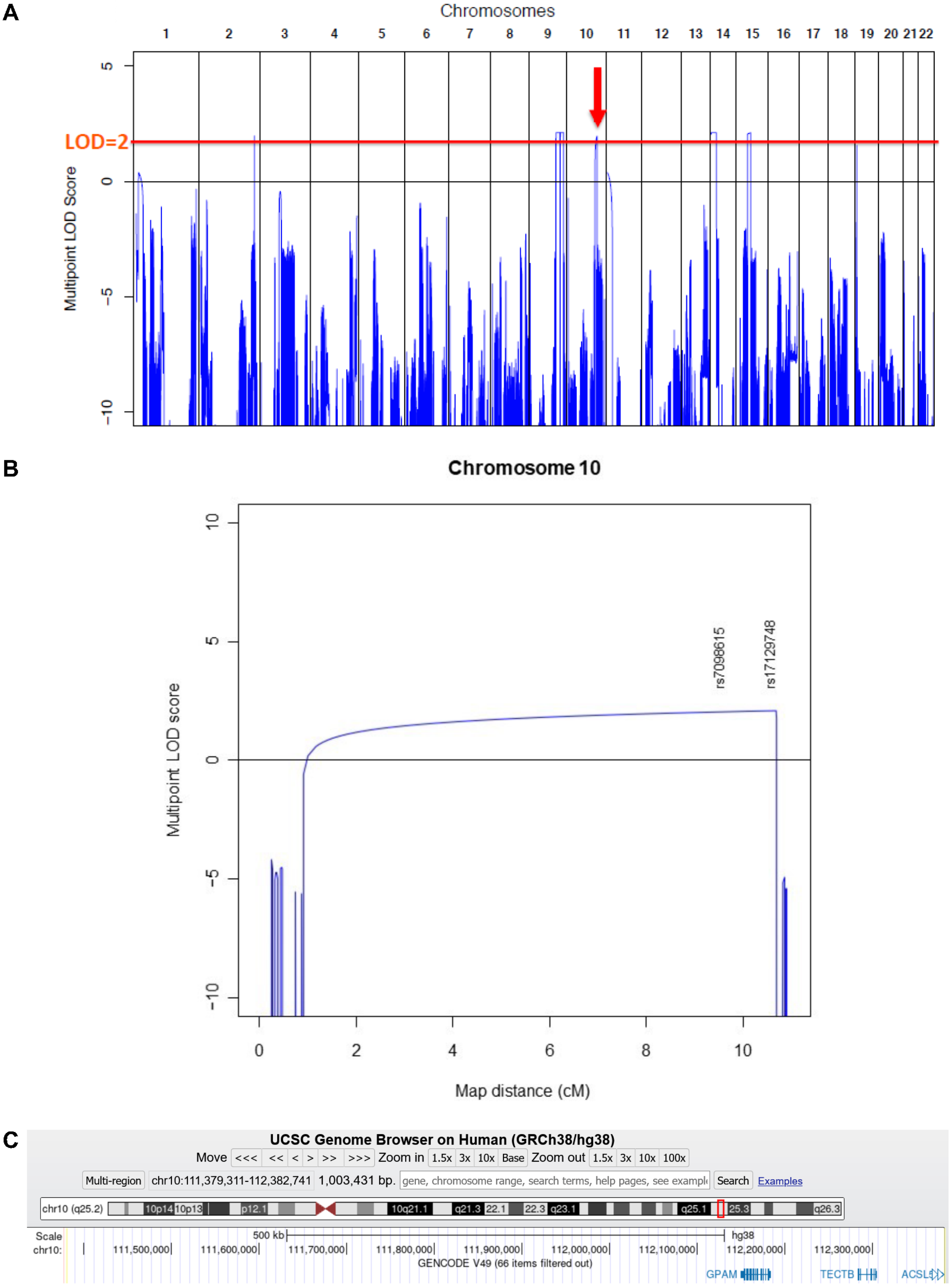


**Supplementary Figure 1. Linkage analysis in Family 1.** (**A**) Linkage analysis showing genome-wide linkage intervals reaching a maximum LOD score >2 (red line). (**B**) Chromosome 10 linkage map showing the significant interval containing *TECTB*. (**C**) Gene content of the chromosomal interval on chromosome 10 reaching LOD significance.

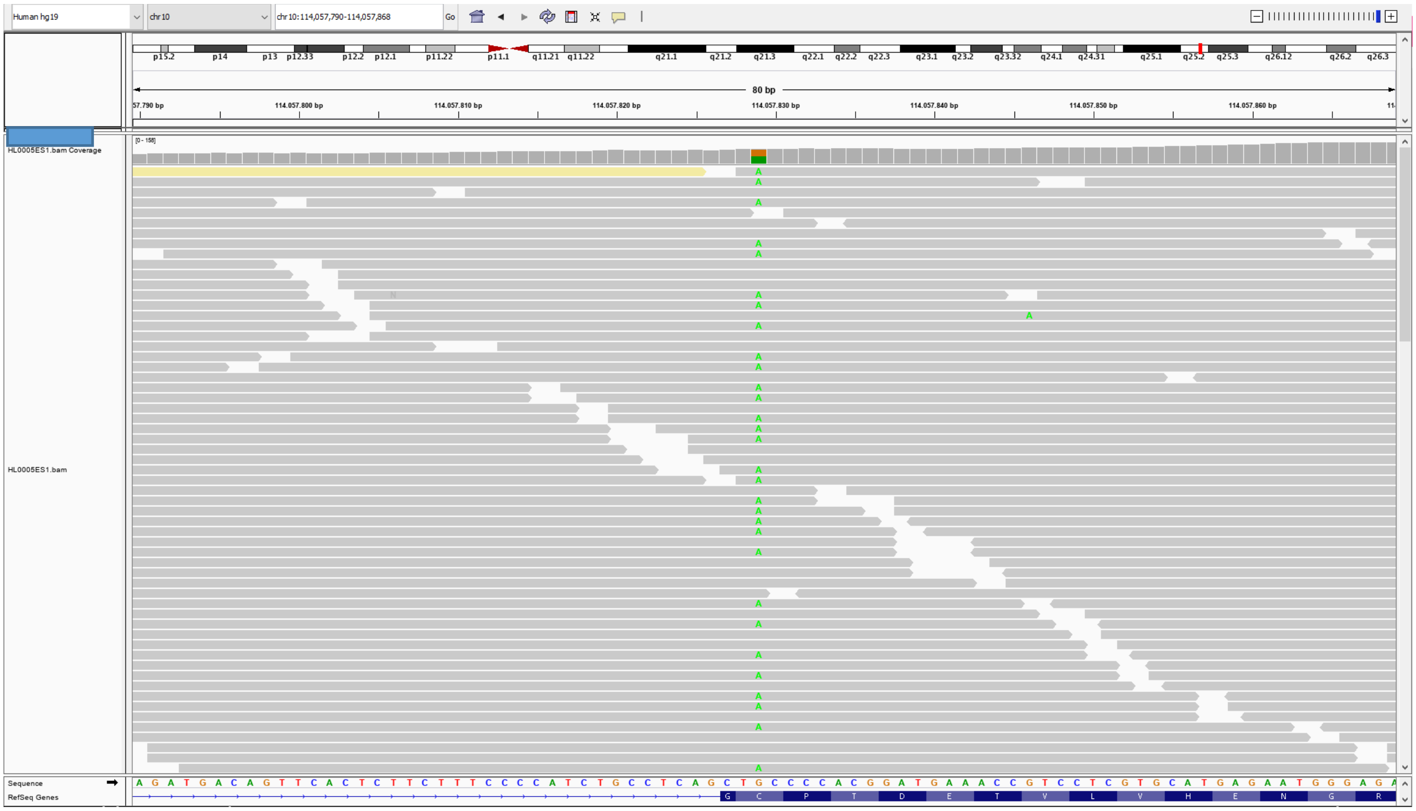

**Supplementary Figure 2. Exome sequencing visualization of the *TECTB* c.674G>A, p.(Cys225Tyr) variant.**

**A**

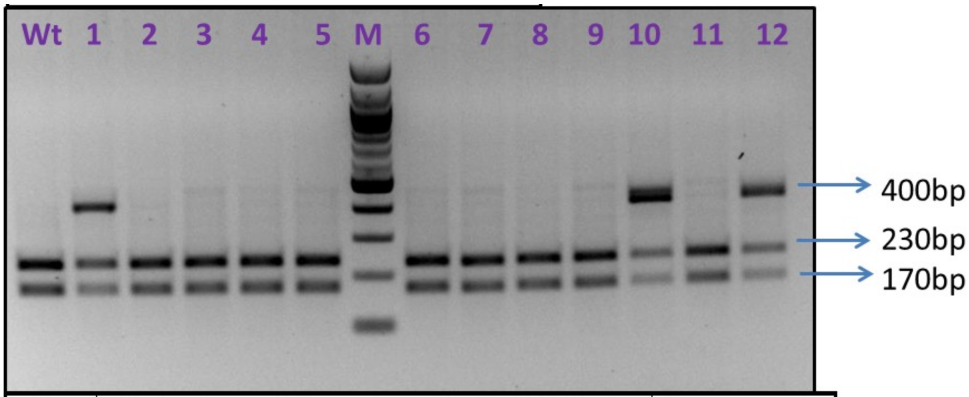


| primers | Product size | Wild type after PvuII digestion | Het after PvuII digestion | Homo mutant after PvuII digestion |
|---|---|---|---|---|
| F1 R1 | 400 | 170 + 230 | 170 + 230 and 400 | 400 |

**B**

Wild type:

C C C A G C T G C C C C A C C G A T

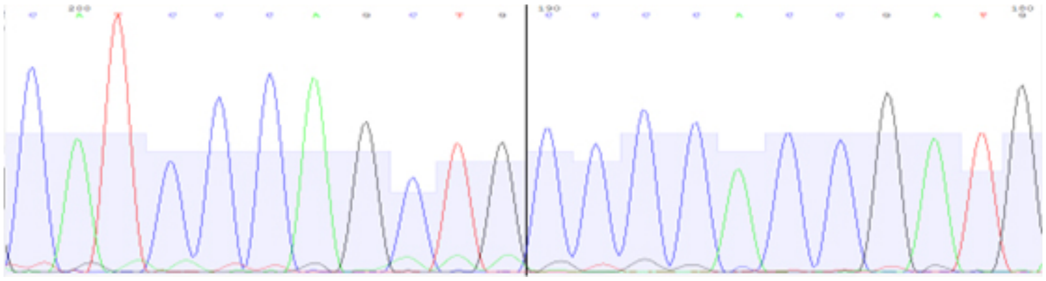

Heterozygous (pup #1):

C C C A G C T G/A C C C C A C C G A T

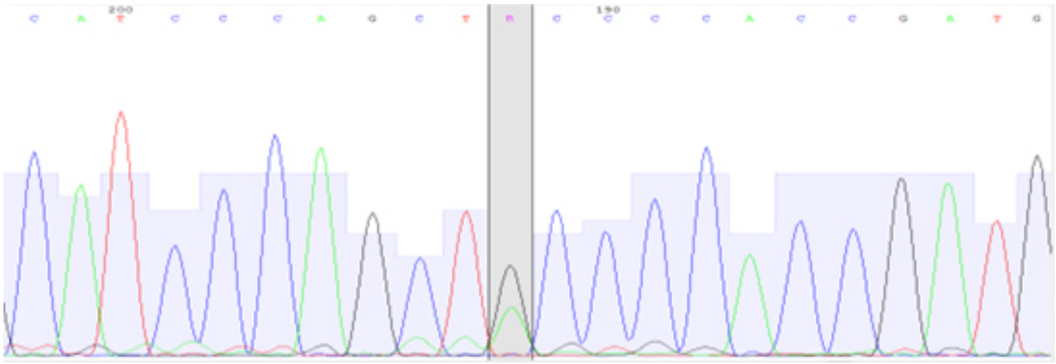

**Supplementary Figure 3. *Tectb-C225Y* mouse line generation. (A)** Genomic DNA extracted from a wild-type mouse and 12 littermate pups born following CRISPR/Cas-9-mediated genome editing was amplified with primers F1 and R1 and digested with *PvuII* as described in the *Material and Methods* section. **(B)** Sanger sequencing of F2R2 PCR products from the wild-type and heterozygous (pup #1) littermates shown above in panel *A*. Sequencing confirmed that pup #1 was correctly modified, while pups #10 and #12 showed evidence of mosaicism.

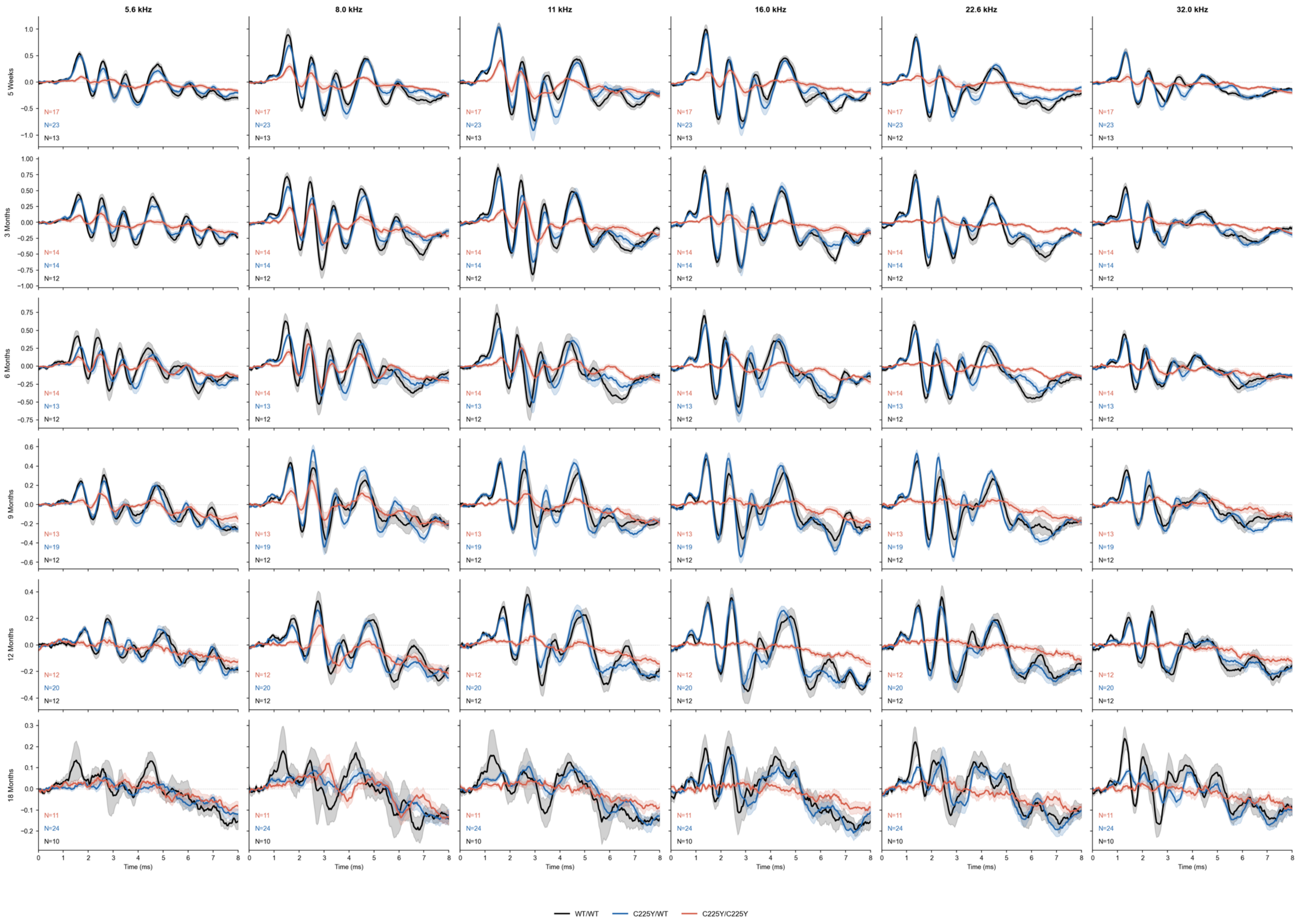


**Supplementary Figure 4. Average ABR Traces Across All Timepoints**. Average ABR traces for wild type, *Tectb*$^{C225Y/+}$, and *Tectb*$^{C225Y/C225Y}$ mice, at 5.6, 8, 11.2, 16, 22.6, and 32 kHz (left to right, 80 dB SPL) at 5 weeks and 3, 6, 9, 12, and 18 months of age (top to bottom).

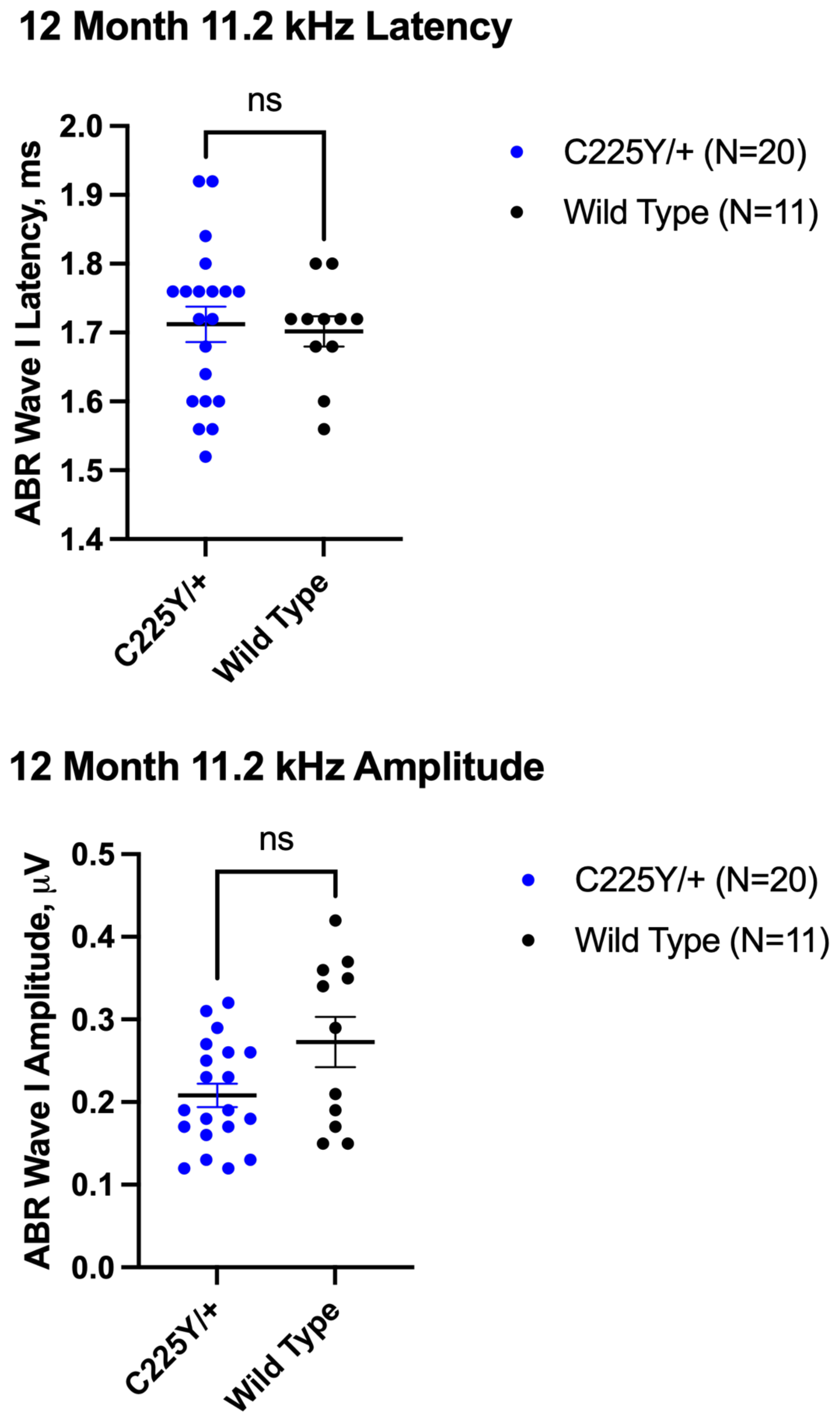


**Supplementary Figure 5. *WT* and *Tectb*$^{C225Y/+}$ mice show no significant differences in ABR wave 1 latency or amplitude values.** ABR wave 1 latency and amplitude measurements in response to 11.2 kHz pure tone stimulation at 90 dB SPL in 12-month-old *Tectb*$^{C225Y/+}$ and wild-type mice. The number of animals per group is indicated within each plot. Data are shown as individual values with the horizontal line representing the mean ± SEM. *Statistical analysis:* Data were first assessed for normality using the Shapiro–Wilk, D'Agostino–Pearson, Anderson–Darling, and Kolmogorov–Smirnov tests. Because both groups were normally distributed, comparisons between genotypes were performed using unpaired two-tailed Welch's t-tests; *ns* – not significant.

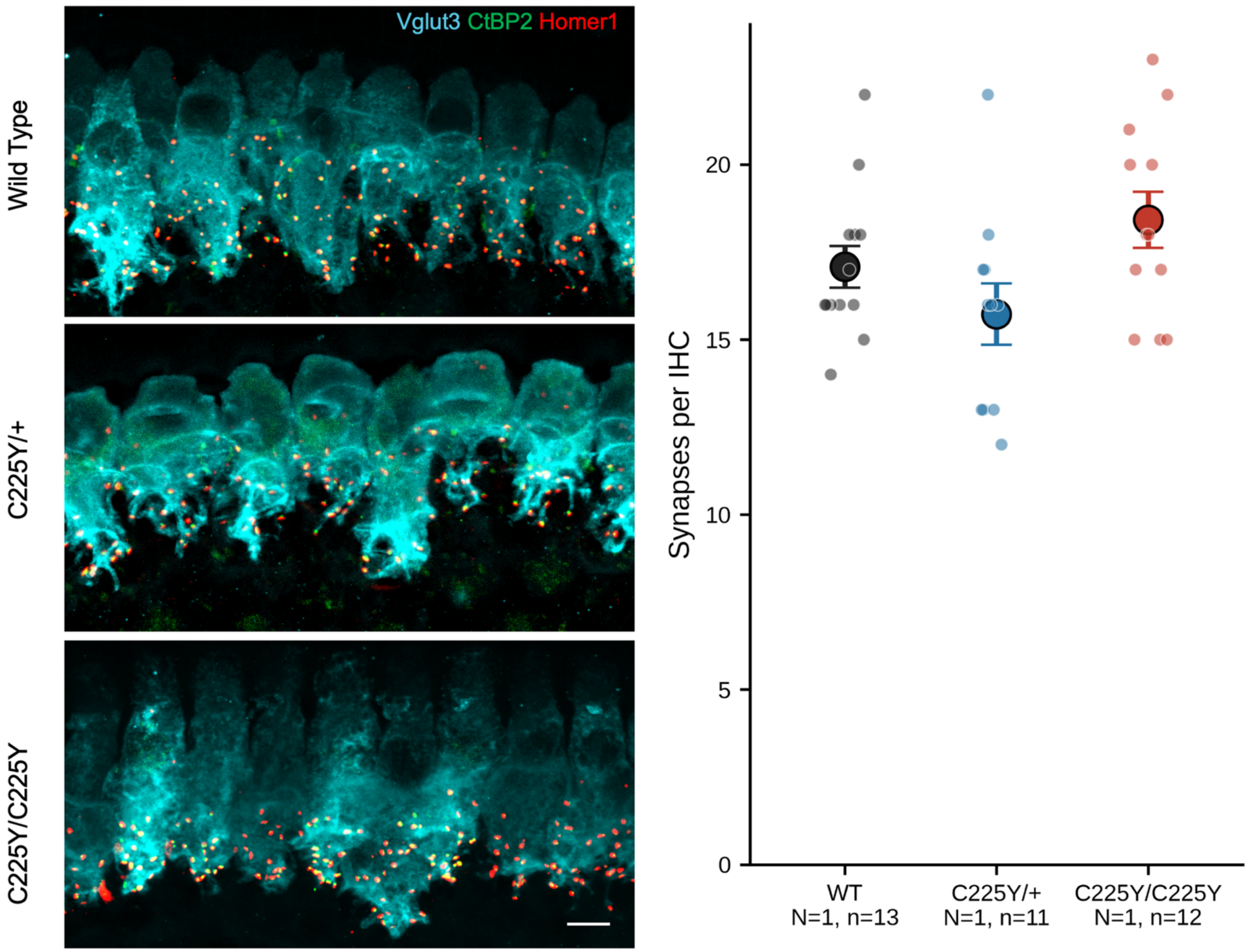


**Supplementary Figure 6. Inner hair cell afferent synapse counts are preserved in *Tectb-C225Y* mice. *Left,*** Representative confocal maximum-intensity projections of the IHC synaptic region in the middle cochlear turn of 10-week-old wild-types (*top*), C225Y/+ (*middle*), and C225Y/C225Y (*bottom*) littermates, immunolabeled for Vglut3 (cyan, IHC cytoplasm), CtBP2 (green, presynaptic ribbons), and Homer1 (red, postsynaptic densities). Scale bar = 5 µm. ***Right,*** Quantification of ribbon synapses per IHC. Synapse counts per IHC were 17.1 ± 0.6 in wild-type, 15.7 ± 0.9 in C225Y/+, and 18.4 ± 0.8 in C225Y/C225Y (N = 1 mouse per genotype, n = 11–13 IHCs; mean ± SEM).

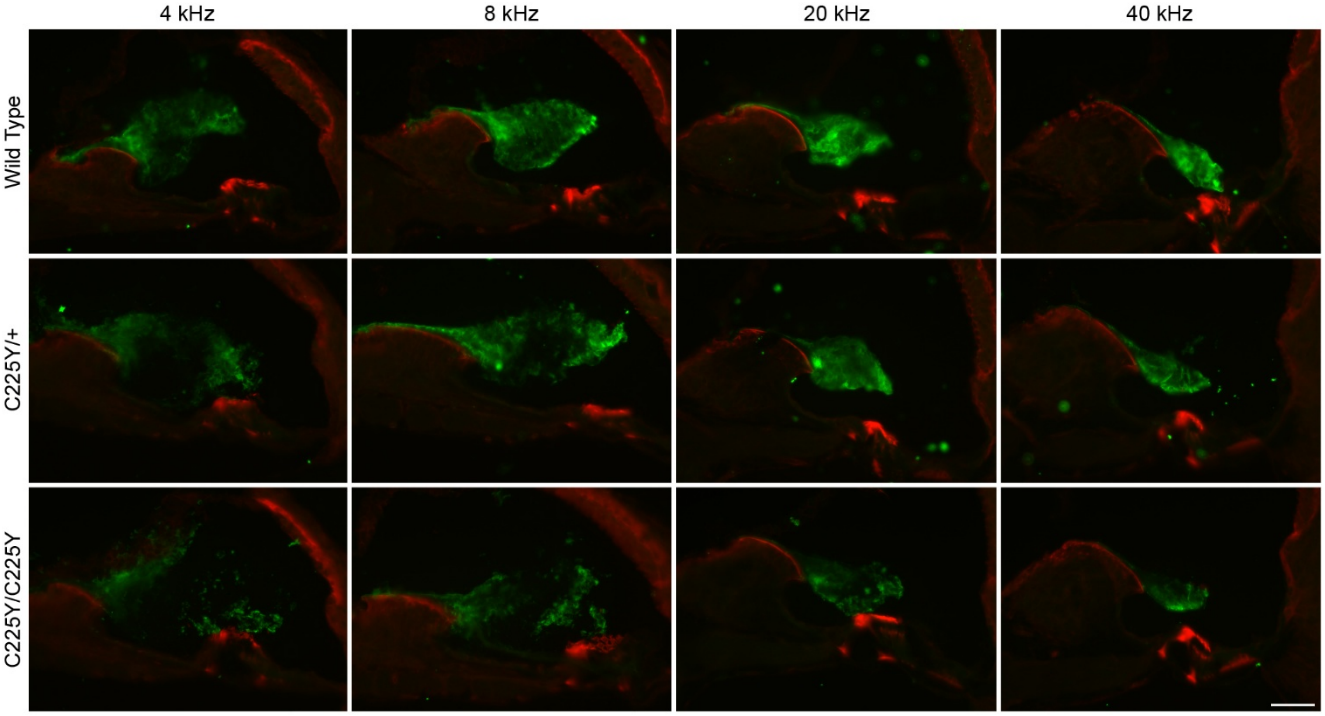


**Supplementary Figure 7. TECTB-C225Y protein is detected within the TM.** Anti-TECTB (R7) immunolabeling of cochlear cryosections from P80 mice, performed in four cochlear locations reveal the presence of anti-TECTB immunoreactivity within the TMs of all three genotypes. These observations indicate that wild-type TECTB and the TECTB-C225Y protein variant are both recognized by the anti-TECTB antibody. *Scale bar* (bottom right), 50 µm.

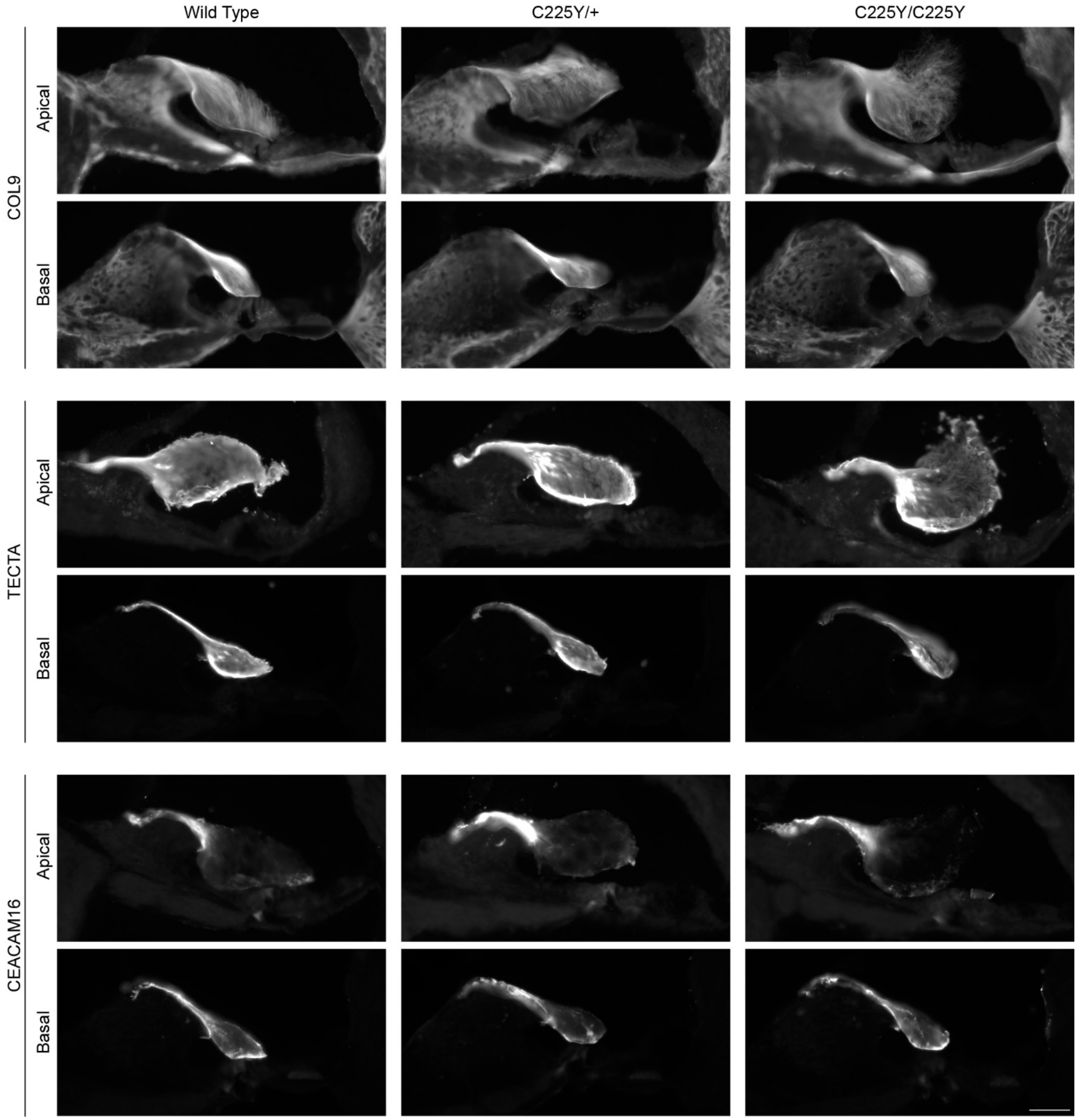


**Supplementary Figure 8. Preserved expression of major tectorial membrane proteins in 6 week old *Tectb-C225Y* mutant mice.** No difference was detected in the expression of other major tectorial membrane proteins, including COL9, TECTA, or CEACAM16, in $\textit{Tectb}^{C225Y/+}$ or $\textit{Tectb}^{C225Y/C225Y}$ mice in either the apical or basal regions of the cochlea. These findings indicate that disruption of tectorial membrane structure in *Tectb-C225Y* mutant mice is not accompanied by loss of these core matrix components, suggesting that the observed phenotype is likely to arise from altered matrix organization rather than changes in major TM protein expression or localization.

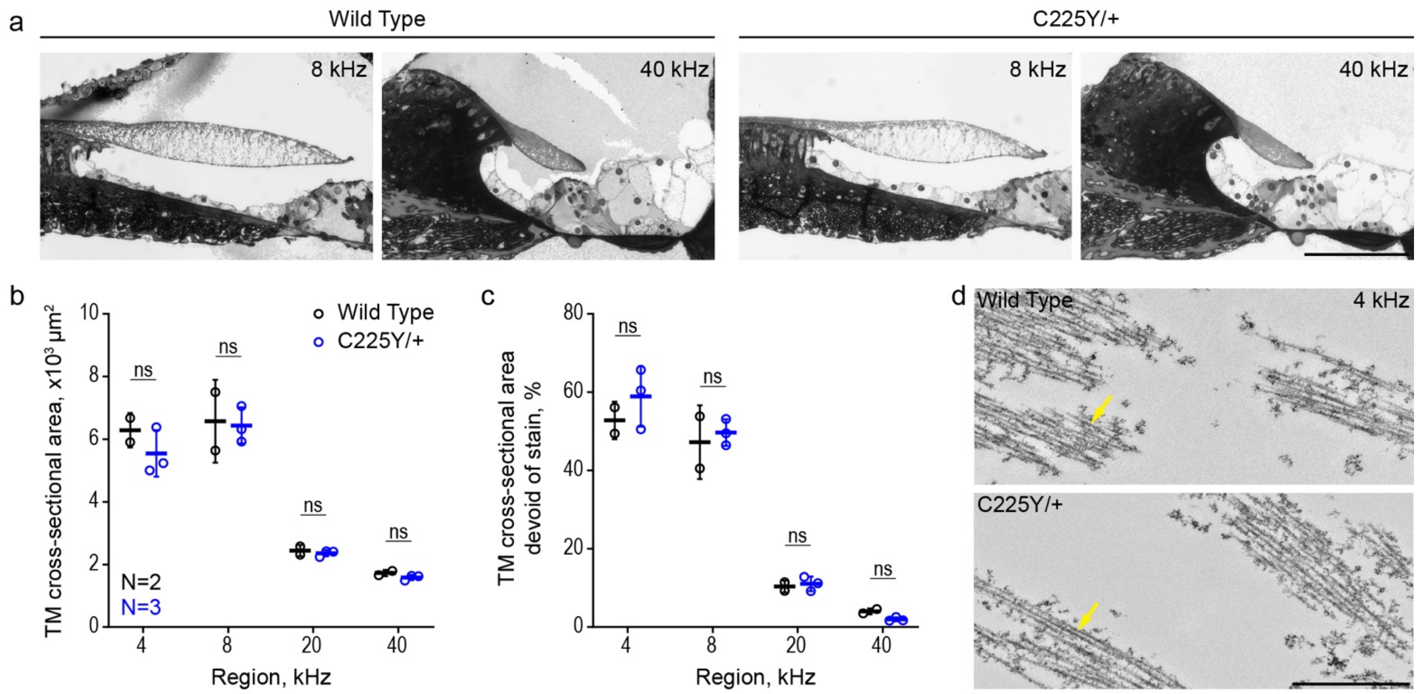


**Supplementary Figure 9, Noise exposure in 18-month-old mice does not alter TM morphology. a,** Toluidine-blue-stained 1 µm sections from the 8 and 40 kHz cochlear regions of wild-type and *Tectb*$^{C225Y/+}$ mice at 18 months of age, fixed 2 weeks after noise exposure. *Scale bar:* 100 µm. **b,** TM Cross-sectional area and **c**, proportion of cross-section area devoid of toluidine blue staining, quantified at 4, 8, 20, and 40 kHz cochlear regions (N=2 wild-type, N=3 *Tectb*$^{C225Y/+}$). No significant differences were detected between genotypes at any region. Data shown as individual values with mean ± SEM. *Statistical analysis*: 2-tailed t test at each frequency with Benjamini-Hochberg correction. Proportions devoid of stain (%) underwent arcsin square root transformation prior to statistical analysis. ns $p > 0.05$. **d,** TEM micrographs of 4 kHz region of the TM. Yellow arrows indicate collagen fibrils, no striated sheet matrix was observed in either genotype, consistent with age-related SSM loss at that age. *Scale bar*: 1 µm.
