## Supplementalry Tables for "*TECTB* Variants Reveal Tectorial Membrane Vulnerability in Dominant Non-Syndromic Hearing Loss"

**Supplementary Table 1**. Summary of known tectorial membrane-expressed genes

| Gene | DFN locus | Inheritance | Human phenotypes | References |
|---|---|---|---|---|
| *CEACAM16* | DFNA4B<br>DFNB113 | AD<br>AR | Progressive mild-to-moderately-severe SNHL<br>Progressive mild-to-moderate SNHL | PMID: 21368133<br>PMID: 29703829 |
| *OTOA* | DFNB22 | AR | Moderate to severe mid-frequency SNHL | PMID: 11972037 |
| *OTOG* | DFNB18B | AR | Mild-to-moderate SNHL | PMID: 23122587 |
| *OTOGL* | DFNB84B | AR | Mild-to-moderate SNHL | PMID: 23122586 |
| *TECTA* | DFNA8/12<br>DFNB21 | AD<br>AR | Moderate to severe, stable or progressive mid-to-high-frequency SNHL; U-shaped audiograms<br>Moderate-to-severe SNHL; flat or U-shaped audiograms | PMID: 9590290<br>PMID: 9949200 |
| *TECTB* | Current study | AD | Variable mild-to-severe SNHL | Current study |

Abbreviations: Autosomal dominant, AD; autosomal recessive, AR; SNHL, sensorineural hearing loss

**Supplementary Table 2**. Risk Factor Questionnaire

| Pedigree ID | Affected Status | Prior Audio | Maternal Infection | Med/Alcohol Exposure | Gentamycin Exposure | Pesticides | Maternal Tobacco | Prematurity | Apnea | Oxygen | Birth Defects | Visual Problems | Hematuria | Neck Mass | Otitis Media | Syncope | Balance Problems | Otorrhea | Treated for Otitis Media | Noise Exposure (TTS) | Occupational Noise |
|---|---|---|---|---|---|---|---|---|---|---|---|---|---|---|---|---|---|---|---|---|---|
| II:1 | Unaffected | 0 | 0 | 0 | 0 | 0 | 0 | 0 | 0 | 0 | 0 | 1 | 0 | 0 | 0 | 0 | 0 | 0 | 0 | Never | 0 |
| II:2 | Unaffected | 0 | 0 | 0 | 0 | 0 | 0 | 0 | 0 | 0 | 0 | 0 | 0 | 0 | 0 | 0 | 0 | 0 | 0 | Never | 0 |
| II:3 | Unaffected | 1 | 0 | 0 | 0 | 1 | 0 | 0 | 0 | 0 | 0 | 1 | 0 | 0 | 0 | 0 | 0 | 0 | 0 | Never | 0 |
| II:4 | Affected | 1 | NA | 0 | NA | 0 | 1 | 0 | NA | NA | 0 | 0 | 0 | 0 | 0 | 0 | 1 | 0 | 0 | Sometimes | 0 |
| II:5 | Affected | 1 | NA | 0 | 0 | 1 | 0 | 0 | 0 | 0 | 0 | 0 | 0 | 1 | 1 | 0 | 1 | 0 | 1 | Never | 0 |
| III:1 | Unaffected | 1 | 0 | 0 | 0 | 1 | 0 | 1 | 0 | 0 | 0 | 0 | 0 | 0 | 0 | 0 | 0 | 0 | 0 | Never | 1 |
| III:2 | Affected | 1 | 0 | 0 | 0 | 1 | 0 | 0 | 0 | 0 | 0 | 0 | 0 | 0 | 0 | 0 | 0 | 0 | 0 | Never | 0 |
| III:3 | Affected | 1 | 0 | 0 | 0 | 0 | 0 | 0 | 0 | 0 | 0 | 0 | 0 | 0 | 0 | 1 | 1 | 0 | 0 | Never | 0 |
| III:5 | Affected | 0 | 0 | 0 | 0 | 1 | 0 | 0 | 0 | 0 | 0 | 0 | 0 | 0 | 0 | 0 | 0 | 0 | 0 | Never | 0 |
| III:6 | Affected | 1 | 0 | 0 | 0 | 1 | 0 | 0 | 0 | 0 | 0 | 0 | 0 | 0 | 0 | 0 | 1 | 0 | 0 | Never | 0 |
| III:7 | Unaffected | 0 | 0 | 0 | 0 | 0 | 0 | 0 | 0 | 0 | 1 | 1 | 0 | 0 | 0 | 0 | 0 | 0 | 0 | Sometimes | 0 |
| IV:1 | Affected | 0 | 0 | 0 | 0 | 0 | 0 | 0 | 0 | 0 | 0 | 0 | 0 | 0 | 0 | 0 | 0 | 0 | 0 | *Shorter survey | |
| | Total N | 12.0 | 10.0 | 12.0 | 11.0 | 12.0 | 12.0 | 12.0 | 11.0 | 11.0 | 12.0 | 12.0 | 12.0 | 12.0 | 12.0 | 12.0 | 12.0 | 12.0 | 12.0 | 11.0 | 11.0 |
| | Positive | 7.0 | 0.0 | 0.0 | 0.0 | 6.0 | 1.0 | 1.0 | 0.0 | 0.0 | 1.0 | 3.0 | 0.0 | 1.0 | 1.0 | 1.0 | 4.0 | 0.0 | 1.0 | 2.0 | 1.0 |
| | Percentage | 58.3 | 0.0 | 0.0 | 0.0 | 50.0 | 8.3 | 8.3 | 0.0 | 0.0 | 8.3 | 25.0 | 0.0 | 8.3 | 8.3 | 8.3 | 33.3 | 0.0 | 8.3 | 18.2 | 9.1 |

Abbreviations: NA, not answered; TTS, temporary threshold shift

**Supplementary Table 3.** Other variants identified in the exome sequencing data in the proband of Family 1.

| | | | | | | Max. MAF gnomAD (v4.1) | | All-of-Us | | | | | | | ACMG Criteria/ Classification | In linked interval? | |
|---|---|---|---|---|---|---|---|---|---|---|---|---|---|---|---|---|---|
| Gene | Transcript | cDNA (c.) | Protein Change (p.) | Zyg | Read Count (Variant:WT) | AF | AF Population | MAF | SIFT | PolyPhen-2 | FATHMM | Mutation Taster | REVEL | CADD | | | Exclusion criteria |
| *BCS1L* | NM_001371449.1 | c.670C>T | p.(Arg224Cys) | Het | 520 (226:294) | 3.1e-5 | European (non-Finnish) | 8.0e-6 | 0.00[1] | 1.0[1] | -2.61[1] | 0.99[1] | 0.778[1] | 32[1] | PM2_P, PP3_M/ VUS | No | Not in linked interval; Gene is associated with recessive disorders |
| *DIS3L2* | NM_152383.5 | c.2074A>C | p.(Asn692His) | Het | 119 (63:56) | 2.2e-5 | European (non-Finnish) | Not present | 0.03[1] | 0.994[1] | 0.87[3] | 0.33[3] | 0.433[2] | 28.6[1] | PM2_P/VUS | No | Not in linked interval; Gene is associated with a recessive disorder |
| *GLYR1* | NM_032569.4 | c.1141C>T | p.(Arg381Cys) | Het | 67 (29:38) | 3.4e-6 | European (non-Finnish) | 4.0e-6 | 0.00[1] | 1.0[1] | -0.24[2] | 0.80[1] | 0.67[1] | 32.0[1] | PM2_P, PP3_P/ VUS | No | Not in linked interval; IMPC mouse with normal hearing |
| *ITIH2* | NM_002216.3 | c.1223T>C | p.(Leu408Ser) | Het | 58 (29:29) | 2.2e-5 | East Asian | 4.0e-6 | 0.03[1] | 1.0[1] | -1.49[2] | 0.39[3] | 0.71[1] | 25.4[1] | PM2_P, PP3_P/ VUS | No | Not in linked interval |
| *PLCB2* | NM_004573.3 | c.431G>T | p.(Arg144Leu) | Het | 201 (115:86) | 1.7e-4 | Admixed American | 2.0e-6 | 0.02[1] | 0.954[1] | 0.93[3] | 0.59[3] | 0.30[2] | 26.8[1] | PM2_P/VUS | No | Not in linked interval |
| *SLC10A2* | NM_000452.3 | c.724_725insC | p.(Phe242Serfs*5) | Het | 44 (19:25) | 0 | -- | 0 | -- | -- | -- | -- | -- | -- | PM2_P/VUS | No | Not in linked interval; Gene is associated with a recessive disorder |
| *SMARCA4* | NM_001387283.1 | c.3355C>T | p.(Arg1119Cys) | Hom | 725 (725:0) | 8.5e-7 | European (non-Finnish) | 0 | 0.00[1] | 1.0[1] | -1.07[2] | 0.99[1] | 0.89[1] | 31[1] | PM2_P, PP3_M/ VUS | No | Not in linked interval; Gene is associated with a dominant disorder, phenotype mismatch |
| *STON1* | NM_006873.4 | c.2167G>A | p.(Gly723Arg) | Het | 51 (18:33) | 0 | -- | -0 | 0.01[1] | 1.0[1] | 2.94[3] | 0.19[3] | 0.30[2] | 31[1] | PM2_P/VUS | No | Not in linked interval; |
| *SUPV3L1** | NM_003171.5 | c.1925G>A | p.(Ser642Asn) | Het | 48 (29:19) | 5.0e-5 | Admixed American | 2.2e-5 | 0.0[1] | 1.0[1] | 1.03[3] | 0.93[1] | 0.47[2] | 35[1] | PM2_P | No | Not in linked interval; |
| *TRPV2* | NM_016113.5 | c.1837C>T | p.(Gln613*) | Het | 192 (106:86) | 1.7e-5 | Admixed American | 1.2e-5 | -- | -- | -- | -- | -- | 37[1] | PM2_P | No | Not in linked interval; Gene associated with perception to pain caused by heat |
| *UBXN2B* | NM_001077619.2 | c.274G>A | p.(Ala92Thr) | Het | 43 (18:25) | 1.7e-5 | Admixed American | 6.0e-6 | 0.00[1] | 1.0[1] | 0.22[3] | 0.54[3] | 0.55[1] | 27.6[1] | PM2_P/VUS | No | Not in linked interval; IMPC mouse with normal hearing |
| *USP3* | NM_006537.4 | c.11C>G | p.(Pro4Arg) | Het | 167 (94:73) | 0 | -- | 0 | 0.00[1] | 0.45[3] | -1.81[1] | 0.26[3] | 0.63[1] | 23.8[2] | PM2_P, PP3_P/ VUS | No | Not in linked interval; Unknown function |

Pathogenicity is represented as [1]deleterious, [2]neutral, or [3]benign prediction, whereas "--" represents variant not scored.

Abbreviations: IMPC, International Mouse Phenotyping Consortium; VUS, variant of uncertain significance; Zyg, zygosity

*Predicted to cause aberrant splicing

**Supplementary Table 4**. List of genes included on the *OtoGenome* hearing loss gene panel

| Gene | OMIM # | Gene | OMIM # | Gene | OMIM # | Gene | OMIM # |
|---|---|---|---|---|---|---|---|
| *ACTG1* | 102560 | *GJB2* | 121011 | *MTTS1* | 590080 | *SLC17A8* | 607557 |
| *ADGRV1* | 602851 | *GJB3* | 603324 | *MYH14* | 608568 | *SLC26A4* | 605646 |
| *ATP6V1B1* | 192132 | *GJB6* | 604418 | *MYH9* | 160775 | *SLC26A5* | 604943 |
| *BSND* | 606412 | *GPSM2* | 609245 | *MYO15A* | 602666 | *STRC* | 606440 |
| *CCDC50* | 611051 | *GRHL2* | 608576 | *MYO1A* | 601478 | *TECTA* | 602574 |
| *CDH23* | 605516 | *GRXCR1* | 613283 | *MYO3A* | 606808 | *TIMM8A* | 300356 |
| *CLDN14* | 605608 | *HGF* | 142409 | *MYO6* | 600970 | *TJP2* | 607709 |
| *CLRN1* | 606397 | *ILDR1* | 609739 | *MYO7A* | 276903 | *TMC1* | 606706 |
| *COCH* | 603196 | *KCNE1* | 176261 | *OTOA* | 607038 | *TMIE* | 607237 |
| *COL11A2* | 120290 | *KCNQ1* | 607542 | *OTOF* | 603681 | *TMPRSS3* | 605511 |
| *CRYM* | 123740 | *KCNQ4* | 603537 | *PCDH15* | 604487 | *TPRN* | 613354 |
| *DFNA5* | 608798 | *LHFPL5* | 609427 | *PDZD7* | 612971 | *TRIOBP* | 609761 |
| *DFNB31* | 607084 | *LOXHD1* | 613072 | *PJVK* | 610219 | *USH1C* | 605242 |
| *DIAPH1* | 602121 | *LRTOMT* | 612414 | *POU3F4* | 300039 | *USH1G* | 607696 |
| *ESPN* | 606351 | *MARVELD2* | 610572 | *POU4F3* | 602460 | *USH2A* | 608400 |
| *ESRRB* | 602167 | *MIR183* | 611608 | *PRPS1* | 311850 | *WFS1* | 606201 |
| *EYA1* | 601653 | *MIR96* | 611606 | *PTPRQ* | 603317 | | |
| *EYA4* | 603550 | *MSRB3* | 613719 | *RDX* | 179410 | | |
| *GIPC3* | 608792 | *MTRNR1* | 561000 | *SERPINB6* | 173321 | | |
